# Blood proteomics of menopause map to brain aging and dementia risk

**DOI:** 10.64898/2026.02.09.26345907

**Authors:** Madeline Wood Alexander, Jennifer S. Rabin, Michelle Caunca, Allesandra Iadipaolo, Louisa Cornelis, Ria Warrier, Keenan A. Walker, Nina Miolane, Veronica Augustina Bot, Brendan Wood, Hamilton Se-Hwee Oh, Tony Wyss-Coray, Albert Pham, Julia Borger, Valentina Diaz, Emily W. Paolillo, Joel Kramer, Laura Pritschet, Caitlin Taylor, Matthew S. Panizzon, Ramiro Eduardo Rea Reyes, Marisa N. Denkinger, Nicholas J. Ashton, Sterling C. Johnson, Emily G. Jacobs, Rowan Saloner, Kaitlin B. Casaletto

**Affiliations:** Hurvitz Brain Sciences Program, Sunnybrook Research Institute, Toronto, Ontario, Canada, M4N 3M5; Rehabilitation Sciences Institute, Temerty Faculty of Medicine, University of Toronto, Toronto, Ontario, Canada, M5G 1V7; Harquail Centre for Neuromodulation, Sunnybrook Health Sciences Centre, University of Toronto, Toronto, Ontario, Canada, M4N 3M5; Division of Neurology, Department of Medicine, Sunnybrook Health Sciences Centre, University of Toronto, Toronto, Ontario, Canada, M4N 3M5; Neurovascular Division, Department of Neurology, Weill Institute for Neurosciences, University of California, San Francisco, California, USA, 94158; Department of Psychological & Brain Sciences, University of California, Santa Barbara, Santa Barbara, California, USA, 93106; Ann S. Bowers Women’s Brain Health Initiative, University of California, California, USA, 93106; Neuroscience Research Institute, University of California, Santa Barbara, Santa Barbara, California, USA, 93106; Department of Physics, University of California, Broida Hall, Santa Barbara, CA, USA, 93106; Laboratory of Behavioral Neuroscience, National Institute on Aging, Baltimore, MD, USA; The Phil and Penny Knight Initiative for Brain Resilience, Stanford University, Stanford, CA, USA; Wu Tsai Neurosciences Institute, Stanford University, Stanford, CA, USA; Graduate Program in Bioengineering, Stanford University, Stanford, CA, USA; Department of Medical Biophysics, University of Toronto, Toronto, ON, Canada; Nash Family Department of Neuroscience, Icahn School of Medicine at Mount Sinai, New York, NY, USA; Brain and Body Research Center of the Friedman Brain Institute, Icahn School of Medicine at Mount Sinai, New York, NY, USA; Department of Genetics and Genomic Sciences, Icahn School of Medicine at Mount Sinai, New York, NY, USA; Ronald M. Loeb Center for Alzheimer’s Disease, Icahn School of Medicine at Mount Sinai, New York, NY, USA; Department of Neurology and Neurological Sciences, Stanford University School of Medicine, CA, USA; Edward and Pearl Fein Memory and Aging Center, Department of Neurology, Weill Institute for Neurosciences, University of California, San Francisco, California, USA, 94158; Department of Psychiatry, University of Pennsylvania, Philadelphia, PA, USA, 19104; Center for Behavior Genetics of Aging, School of Medicine, University of California, San Diego, La Jolla, CA, USA, 92093; Wisconsin Alzheimer’s Disease Research Center, School of Medicine and Public Health, University of Wisconsin, Madison, WI, USA, 53792-2420; Banner Sun Health Research Institute, Sun City, AZ, USA, 85351; Banner Alzheimer’s Institute, Phoenix, AZ, USA, 85006; Wisconsin Alzheimer’s Institute, School of Medicine and Public Health, University of Wisconsin, Madison, WI, USA, 63726

## Abstract

Menopause is a hallmark process in biological aging that has been implicated in later neurodegenerative risk. We leveraged proteomics data from multiple cohorts to identify biological changes underlying menopause and its links to brain aging. In *N*=80 rigorously staged (STRAW+10) pre-, peri-, and postmenopausal women with serum NULISAseq proteomics, spontaneous menopause was characterized by dysregulation in inflammatory, synaptic, metabolic, and Alzheimer’s disease biologic processes, which tracked more strongly with hormones than with age. Validation analyses in age-matched pre/peri-versus postmenopausal women with plasma Olink proteomics (*N*=2,814) replicated the observed proteomic shifts and revealed broader menopause-related upregulation of inflammatory and catabolic processes plus accelerated organ and cell aging, including brain aging. In four independent cohorts of older women (average age=60.7 to 72.1 years; total *N*=11,925), menopause-related proteomic elevations consistently associated with poorer cognitive outcomes. The molecular signatures of menopause may inform biomarkers or therapeutic targets for brain health in midlife women.

## INTRODUCTION

Accumulating data point towards menopause as a critical inflection point in the aging process.^1^ Universal to all midlife individuals with ovaries, menopause is characterized by a steep decline in ovarian hormones (17β-estradiol, progesterone) and a sustained rise in follicle-stimulating hormone (FSH). These endocrine shifts are associated with widespread multi-system health effects^1–4^ that may be relevant for brain aging and dementia risk.^5–9^ Canonical menopause symptoms, including vasomotor (hot flashes/night sweats), sleep problems, and mood/cognitive changes, are neurologic in origin, suggesting an acute neural response to the changing endocrine environment.^10–12^ A recent review estimates 26-95% of women experience cognitive symptoms during menopause (word-finding, attentional difficulties), and finds that overall observational studies show subtle performance declines (especially in verbal learning and memory) that fall within normative expectations over the menopause transition.^12^ Further, experimental data suggest that hypothalamic aging may precede and can causally trigger ovarian senescence, inducing menopause.^13,14^ Together, these findings strongly implicate brain involvement in the menopause cascade.

Menopause may represent a midlife window to understand and intervene on incipient brain aging and dementia risk. Although dementia symptoms typically emerge in the mid-70s, biomarker evidence strongly suggests that the pathophysiology of Alzheimer’s disease (AD) begins decades earlier,^15,16^ placing menopause (average age=51^17^) in the coincident timeframe for disease pathogenesis. Increasing evidence has identified earlier and surgical menopause as risk factors for AD.^5–8,18^ Additionally, vasomotor symptoms associate with poorer memory performance, white matter hyperintensities, and AD biomarkers–hallmarks of early dementia risk.^19–21^ These data position menopause as a neurologic transition that offers a framework for understanding early manifestations of brain aging and identifying changes that may harken later-life dementia risk in women.

Despite these emerging links, human research investigating the biological pathways by which menopause influences brain aging is scarce. Limited existing proteomics studies of menopause have identified changes in targets involved in metabolic, vascular, and immune processes.^22,23^ However, existing studies did not employ rigorous menopause staging or proteomic platforms targeted at brain aging, and no studies have linked the molecular profiles of menopause to cognitive outcomes.

We characterized blood-based biological signals associated with spontaneous menopause in healthy midlife women with a restricted age range (43–58) and rigorous menopause staging.^24^ Using ultrasensitive NUcleic acid Linked Immuno-Sandwich Assay (NULISAseq) proteomics,^25^ we showed that menopause associated with shifts in inflammatory, synaptic, metabolic, and AD biologic processes. These signals tracked more strongly with sex hormones than with age. Pro-inflammatory proteins were especially upregulated in women with vasomotor symptoms. Validation in a large sample of age-matched pre/peri-versus postmenopausal women with plasma Olink proteomics replicated menopause-associated proteomic shifts and showed broader upregulation of inflammatory and catabolic processes, including accelerations in multisystem organ and cellular aging. Across four cohorts of older women with average ages of 61–72 years, menopause-related proteomic elevations consistently associated with worse cognitive outcomes. Together, these findings highlight reproducible signatures of menopause that overlap with molecular drivers of later life brain health, underscoring menopause as a discovery framework for understanding the earliest manifestations of brain aging and risk and resilience to dementia.

## RESULTS

### Spontaneous menopause cohort

We included *N*=30 premenopausal, *N*=26 perimenopausal, and *N*=24 postmenopausal women, plus *N*=36 age-matched men for comparison (Tab.S1; Fig.1A). In women, age ranged from 43–58, and age differences by menopause stage were statistically significant but small (Fig.1B). The vast majority were late premenopausal (STRAW+10 stages -3b/-3a), perimenopausal (−2/-1), or early postmenopausal (1a/1b/1c; *N*=1 postmenopausal woman stage 2; *N*=1 perimenopausal woman for whom exact stage could not be ascertained). There were no significant age differences by sex and no menopause group differences in body mass index. Where not included in the text, statistics are presented in Extended Data.

**Figure 1.**
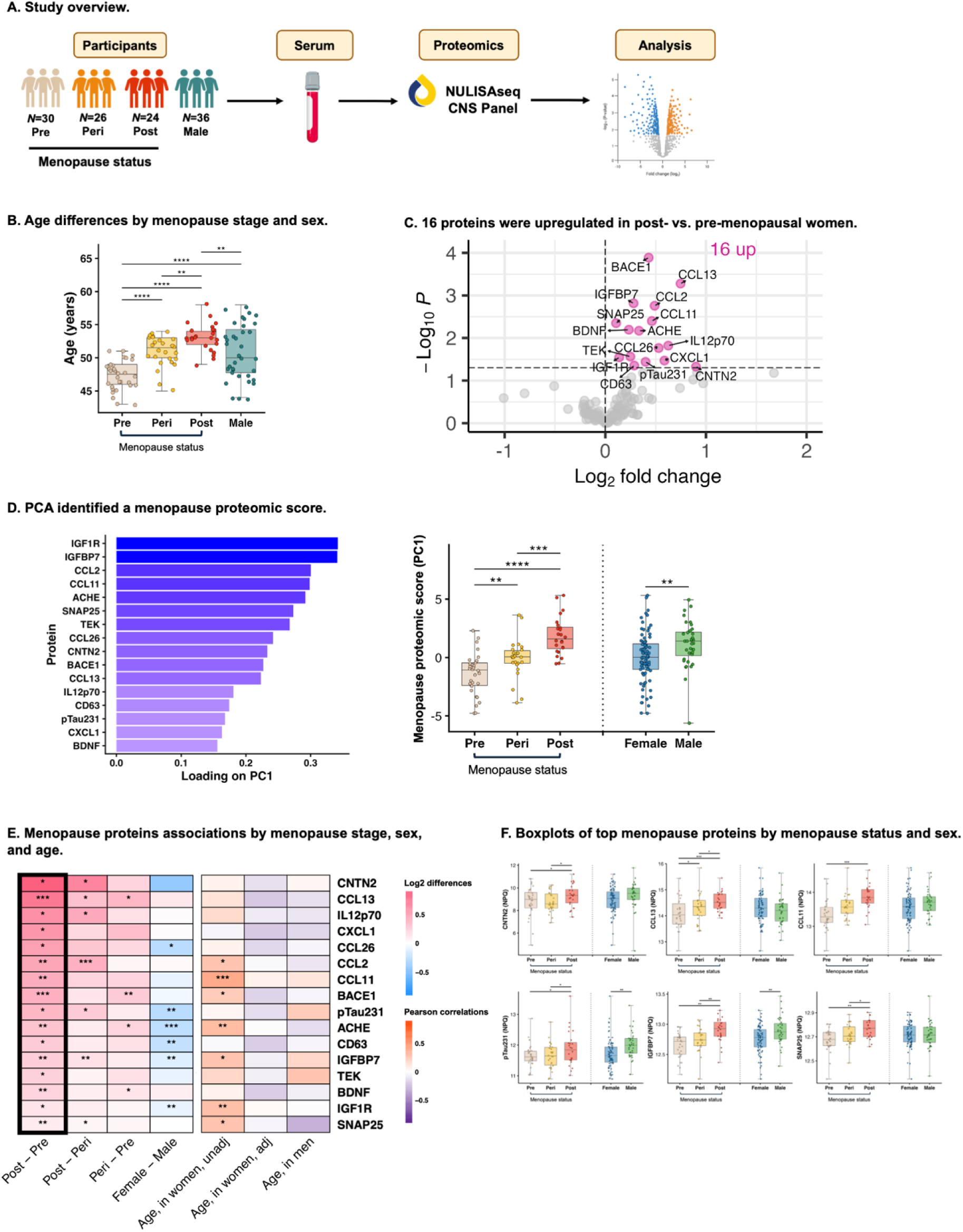
Blood-based proteomics of spontaneous menopause. **A.** Blood was collected in 30 premenopausal, 26 perimenopausal, and 24 postmenopausal women, plus 36 men for comparison. Serum was analyzed for 118 proteins via NUcleic acid Linked Immuno-Sandwich Assay (NULISAseq CNS panel). Age-adjusted analyses identified differences in protein levels across menopause stages and sex. **B.** Participant ages ranged from 43 to 58 years. Despite significant group differences in expected directions, menopause stages exhibited substantial overlap in age. **C.** Analysis of variance (ANOVA) revealed 16 proteins that were significantly upregulated in postmenopausal women relative to premenopausal women, reflecting immune, synaptic, metabolic, and neurodegenerative biology. **D.** Principal components analysis (PCA) performed on these 16 proteins identified a menopause proteomic score (i.e., PC1) that increased across menopause stages. **E.** In age-adjusted analyses, many of the 16 menopause proteins showed significant elevations across menopause stages (i.e., pre-, to peri-, to post), and some showed sex differences. Several proteins significantly correlated to age in women in bivariate analyses, but these associations with age attenuated and reversed direction after accounting for menopause status. No proteins significantly correlated to age in men. **F.** Boxplots of top menopause proteins revealed stepwise increases across the menopause transition. \**p*<.05, \*\**p*<.01, \*\*\**p*<.001.

#### Inflammatory, synaptic, and neuropathology-related proteins were elevated in postmenopausal women

We first examined differential serum protein abundance in postmenopausal vs. premenopausal women (Fig.1C). Adjusting for age, 16 proteins were elevated in postmenopausal women, reflecting inflammatory (CCL13, IL-12p70, CXCL1, CCL26, CCL2, CCL11, TEK, CD63), synaptic/neuronal network (CNTN2, ACHE, SNAP-25, BDNF), metabolic (IGFBP7, IGF1R), and AD (BACE1, p-tau231) biology. To reduce data dimensionality and noise, more holistically capture biology represented, and mitigate multiple comparisons, we performed principal components analysis (PCA) on these 16 targets to estimate a menopause proteomic score that summarizes these top menopause-related changes. The first principal component (PC1) explained 28.7% of the variance in menopause proteins (Fig.S1), whereby higher values reflected a more postmenopausal-like proteome (Fig.1D). Confirming this, age-adjusted analyses revealed stepwise increases in PC1 (i.e., the menopause proteomic score) across the menopause transition (post [vs. peri]: β=2.20, 95% CI=1.178, 3.21, *p*<.0001, peri [vs. pre]: β=1.87, 95% CI=0.772, 2.98, *p*=.001). We assessed stability of PC1 loadings using bootstrap resampling (1000 iterations), which showed low *SD*s (<0.08) and high sign consistency (>0.95) across all proteins (Fig.S2).

In women, age was significantly associated with higher menopause proteomic scores (cor=0.36, *p*=.001). However, this association with age was no longer significant and reversed directionality after adjusting for menopause stage (β=-0.14, 95% CI=-0.31, 0.03, *p*=.11). Unadjusted linear estimates showed a stepwise effect of menopause stage on the proteomic score. Relative to premenopausal, perimenopausal status was associated with a 1.36-unit (*SD*) increase in menopause scores, and postmenopausal status with a 3.25-unit increase. By contrast, each year of age was associated with a 0.24-unit higher menopause score. To assess the potential modulating role of age, we also tested interactions between menopause stage and age on the menopause score, which were not significant (post [vs. pre]*age: β=0.003, 95% CI= 0.44, 0.45, *p*=.99; peri [vs. pre]*age: β=0.17, 95% CI=-0.24, 0.57, *p*=.42). We computed the same composite score in men and found age did not correlate to the menopause score in men (cor=-0.02, *p*=.89). Overall, these findings suggest that the observed proteomic changes were driven more by menopause stage than chronological age in women.

Notably, after accounting for age, p-tau231 was the only canonical AD biomarker (versus p-tau181, AB40, AB42, GFAP, NFL) that significantly differed by menopause stage (Fig.S3). Examining sex differences, men had higher “menopause proteomic scores” than women overall adjusting for age (β=1.13, 95% CI=0.27, 1.90, *p*=.009).

#### Menopause proteins tracked with estradiol and FSH

Estradiol, FSH, and progesterone significantly differed by menopause stage, while testosterone, SHBG, and DHEAS did not (Tab.S1, Figs.2A-B). Therefore, we focused on the former three in proteomic analyses. Age-adjusted models showed the menopause proteomic score inversely associated with estradiol (β=-0.89, 95% CI=-1.37, -0.41, *p*=.0004), positively associated with FSH (β=1.29, 95% CI=0.78, 1.80, *p*<.0001), and did not associate with progesterone (β=-0.23, 95% CI=-0.71, 0.25, *p*=.34; Fig.2C). When simultaneously modeling all three hormones together, only FSH was significantly associated with the menopause score (FSH: β=1.13, 95% CI=0.51, 1.78, *p*=.0005; estradiol: β=-0.43, 95% CI=-0.97, 0.11, *p*=.12; progesterone: β=0.24, 95% CI=-0.20, 0.68, *p*=.28). Furthermore, in the same multivariable model, age was not associated with the menopause score (β=-0.01, 95% CI=-0.17, 0.14, *p*=.88), suggesting that the menopause score is more strongly related to hormone levels than to age when modelled together. Post-hoc analyses examining individual proteins showed distinct hormone-protein relationships such that CCL26, CCL2, CD63, and TEK inversely tracked with 17β-estradiol; CNTN2, CCL11, BACE1, ACHE, and IGFBP7 positively tracked with FSH; and only ACHE tracked with progesterone (Figs.2D-E). A network analysis estimating pairwise correlations at a threshold of r=0.3 illustrated that estradiol-related proteins and FSH-related proteins clustered into two separable groups (Fig.2F), suggesting that different menopause proteins may reflect distinct hormone-related biology.

**Figure 2.**
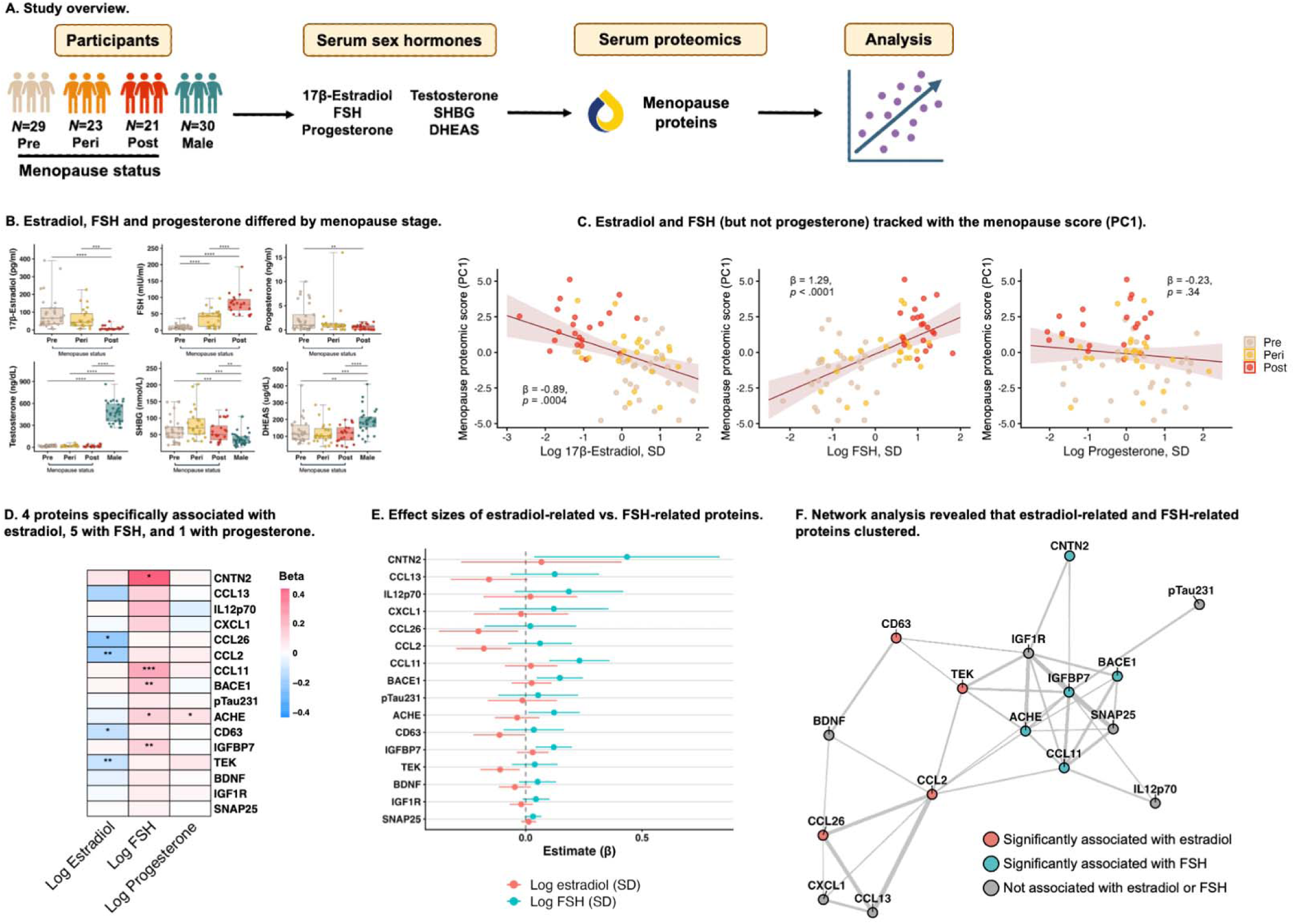
Menopause proteins tracked with estradiol and follicle-stimulating hormone (FSH). **A.** Serum was analyzed for sex hormones: 17β-estradiol, follicle-stimulating hormone (FSH), and progesterone (in women only), plus testosterone, sex-hormone binding globulin (SHBG), and dehydroepiandrosterone sulfate (DHEAS) in all participants. Analyses examined associations between menopause proteins and hormone levels. **B.** Estradiol, FSH, and progesterone differed by menopause stage. **C.** Age-adjusted regression models revealed that estradiol and FSH (but not progesterone) significantly related to the menopause proteomic score. **D.** In models simultaneously testing associations of hormone levels with individual proteins, FSH was uniquely associated with 5 proteins, estradiol was uniquely associated with 4, and progesterone was uniquely associated with 1. **E.** A forest plot of beta coefficients for estradiol and FSH from age- and progesterone-adjusted analyses showed that menopause proteins demonstrate specificity in their associations with sex hormones. CNTN2, CCL11, BACE1, ACHE, and IGFBP7 are predicted only by FSH, and CCL26, CCL2, CD63, and TEK only by estradiol. **F.** Network analysis demonstrated that estradiol-associated (CCL13, CCL26, CCL2, TEK) and FSH-related (CNTN2, CCL11, BACE1, ACHE, IGFBP7) proteins clustered into two separate groups. \**p*<.05, \*\**p*<.01, \*\*\**p*<.001.

#### Women with vasomotor symptoms showed the most pronounced increases in inflammatory menopause proteins

In peri- and postmenopausal women, we tested whether menopause symptoms were associated with the menopause proteomic score (Fig.3A). Night sweats were related to higher (i.e., more postmenopausal) scores (β=0.94, 95% CI=0.00, 1.88, *p*=.05), while vaginal dryness was associated with lower scores (β=-1.15, 95% CI=-2.17, -0.13, *p*=.03). There were no associations between hot flashes or irritability and the menopause score (hot flashes: β=0.43, 95% CI=-0.55, 1.42, *p*=.38; irritability: β=0.32, 95% CI=-0.74, 1.38, *p*=.55). In post hoc analyses, CXCL1, CCL26, CCL13, and CCL2 were upregulated in women with night sweats, while CNTN2, ACHE, and IGFBP7 were downregulated in women with vaginal dryness (Figs.3B-C).

**Figure 3.**
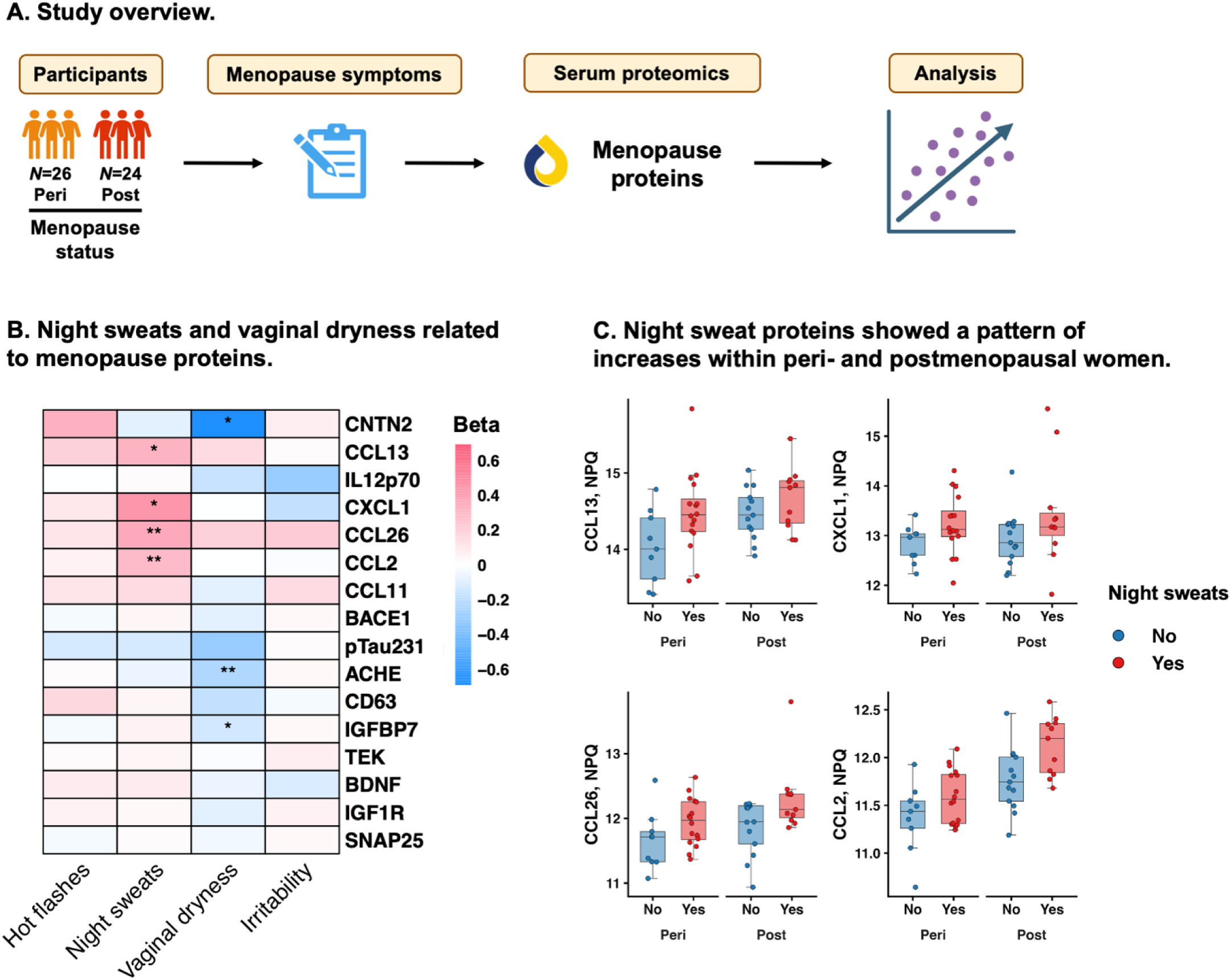
Menopause symptoms associated with menopause protein levels in peri- and postmenopausal women. **A.** Peri- and postmenopausal women self-reported hot flashes, night sweats, vaginal dryness, and irritability. Analyses tested associations of menopause symptoms with menopause proteomic scores and individual proteins, adjusted for age and menopause stage. **B.** Women with night sweats had significantly elevated levels of CCL13, CXCL1, CCL26, and CCL2. Conversely, women with vaginal dryness had significantly lower levels of CNTN2, ACHE, and IGFBP7. **C.** Boxplots of top night sweats proteins showed that women with night sweats had a pattern of elevated protein levels within each menopause stage.

### Cross-platform menopause validation cohort

#### Menopause proteomic shifts replicate in a large independent cohort

In the UK Biobank (UKB) validation cohort including equal groups of age-matched pre/peri- and postmenopausal women aged 45-60 (total *N*=2,814; Tab.S7), 44% of plasma proteins (1,300/2,923 Olink) were differentially abundant (FDR-*p*<.05) by menopause stage (1,146 upregulated in post vs. pre/peri, 154 downregulated in post vs. pre/peri; Fig.4A-C). Top hits included markers of ovarian hormone signaling, including decreased progestogen-associated endometrial protein (PAEP) and increased FSH subunits beta (FSHB) and alpha (CGA; Fig.4D). We assessed whether the specific menopause proteins identified in the UCSB cohort also showed menopause stage differences in the UKB. Of the 13 menopause proteins measured on both proteomic platforms, 9 clearly replicated across cohorts, with effect size concordance (r=0.39) and strong overlap of largest-magnitude hits (Fig.4E). Replication analyses were robust using the menopause proteomic score as the outcome, tested in a smaller subset of UKB women with non-missing values on all 13 available menopause proteins (*N*=1,976; post [vs. pre/peri]: β=0.22, 95% CI=0.13-0.31, *p*<.001).

**Figure 4.**
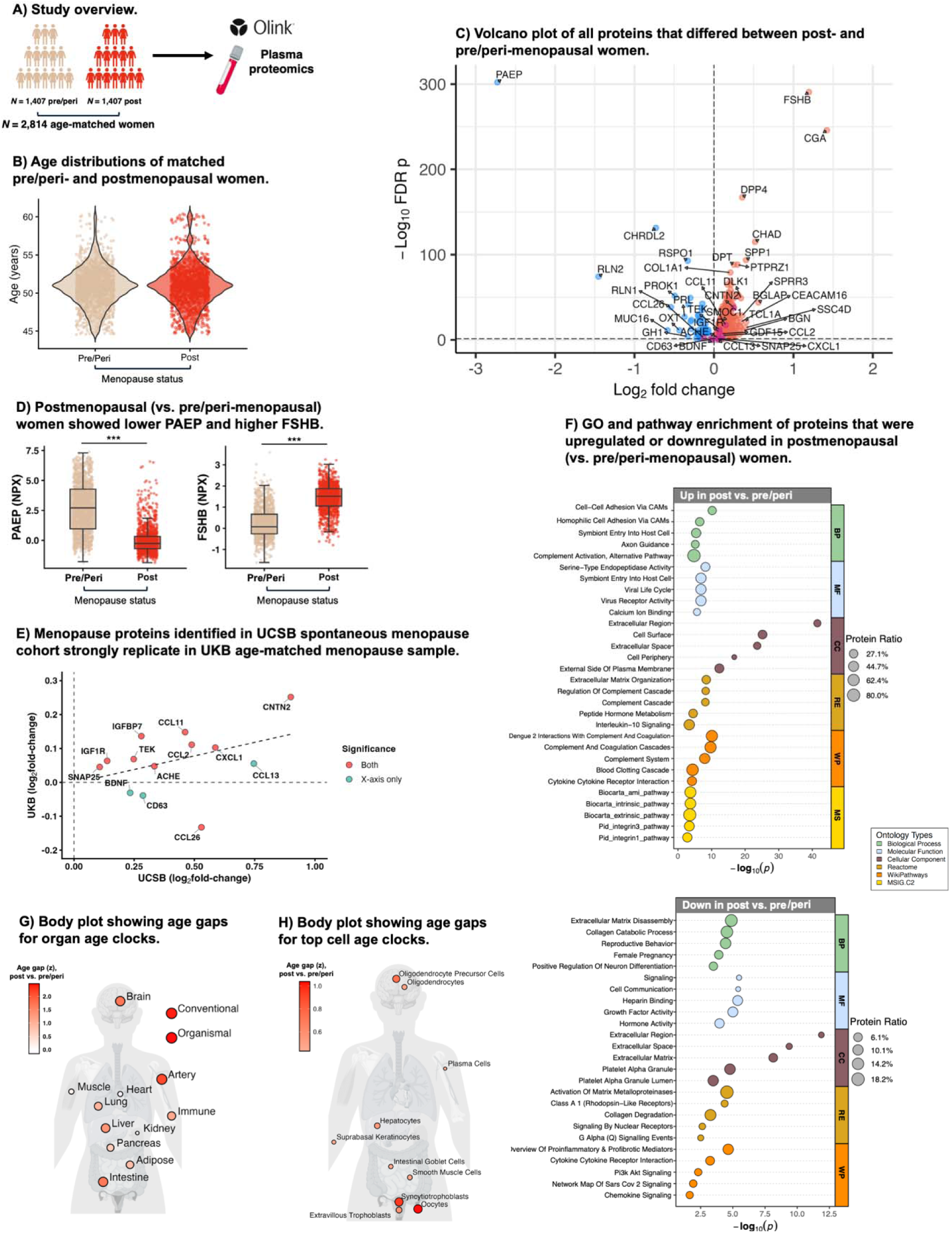
Menopause proteomic shifts replicate cross-platform in age-matched pre- and postmenopausal women in the UK Biobank. **A-B.** Pre- and postmenopausal women aged 45-60 were propensity-matched on age with a 1:1 ratio to form the analytic sample (*N*=2,814). **C.** Age-adjusted differential abundance analyses of Olink proteins demonstrated 1,146 upregulated and 154 downregulated proteins in postmenopausal (vs. premenopausal) women. **D**. Top hits included markers of ovarian hormone signaling, including progestogen-associated endometrial protein (PAEP) and FSH subunits beta (FSHB). **E.** Effect sizes for menopause proteins identified in UCSB demonstrated concordance with those in UKB, showing strong overlap of the largest-magnitude hits in both cohorts. **F**. Gene ontology (GO) and pathway enrichment analysis revealed menopause-associated downregulation of proteins enriched for growth factor and reproductive signaling and upregulation of proteins enriched for cytokine signaling, complement activation, and extracellular matrix catabolism. **G.** Body plot showing organ age associations with menopause stage. **H.** Body plot showing top ten cell aging measures associated with menopause stage. \**p*<.05, \*\**p*<.01, \*\*\**p*<.001.

We performed gene ontology (GO) and pathway analysis on the broader list of 1,300 Olink proteins differentially abundant by menopause stage. These analyses revealed menopause-associated downregulation of proteins enriched for growth factor and reproductive signaling and upregulation of proteins enriched for cytokine signaling, complement activation, and extracellular vesicle–related processes (Fig.4F). Next, in *N*=2,340 age-matched pre/peri- and postmenopausal women, we estimated proteomic aging clocks (Extended Data) and assessed whether menopause stage associated with organ and cell aging. Relative to pre/peri-menopausal women of the same age, in age-adjusted analyses, postmenopausal women had larger (i.e., less favourable) age gaps on 11 of 13 organ age measures (Fig.S4), and 36 of the 38 cell age measures (Fig.S5). Top menopause-associated organ ages included artery, brain and intestine aging, plus conventional and organismal (i.e., overall, organ-shared) aging (Fig.4G). For cell aging, top measures included female reproductive cells (e.g., oocytes, syncytiotrophoblasts), and oligodendrocyte precursor cells, supporting the biological validity of menopause classifications in the UKB and highlighting the potential role of menopause in brain aging (Fig.4H).

### Aging cohorts

#### Menopause symptoms associated with menopause protein levels in later life

In 89 women (mean[*SD*] age=69.2[6.84]) in the Wisconsin Registry for Alzheimer’s Prevention (WRAP; Tab.S10), we examined whether history of menopause symptoms showed associations with the identified menopause proteins years to decades after menopause (Fig.5A). Higher menopause proteomic scores associated with hot flashes (β=0.58, 95% CI=0.06, 1.11, *p*=.03), problems sleeping (β=0.45, 95% CI=0.03, 0.86, *p*=.04), and total symptom count (β=0.16, 95% CI=0.02, 0.30, p=.03; Fig.5B). Associations between other menopause symptoms and the proteomic score were not significant (mood swings: β=0.42, 95% CI=-0.01, 0.85, *p*=.06; night sweats: β=0.04, 95% CI=-0.37, 0.46, *p*=.84; depression: β=0.18, 95% CI=-0.35, 0.71, *p*=.50; sexual dysfunction: β=0.41, 95% CI=-0.23, 1.05, *p*=.21). Post-hoc analyses showed that hot flashes associated with increases in IL12p70 and CCL2; problems sleeping with increases in BACE1 and IGF1R; and total symptom count with increases in IL12p70, BACE1, and IGF1R (Fig.5C). CCL2 was the only individual protein associated with vasomotor symptoms in both older women and midlife women, suggesting a link between vasomotor symptoms and CCL2 that may persist into later life (Fig.5D).

**Figure 5.**
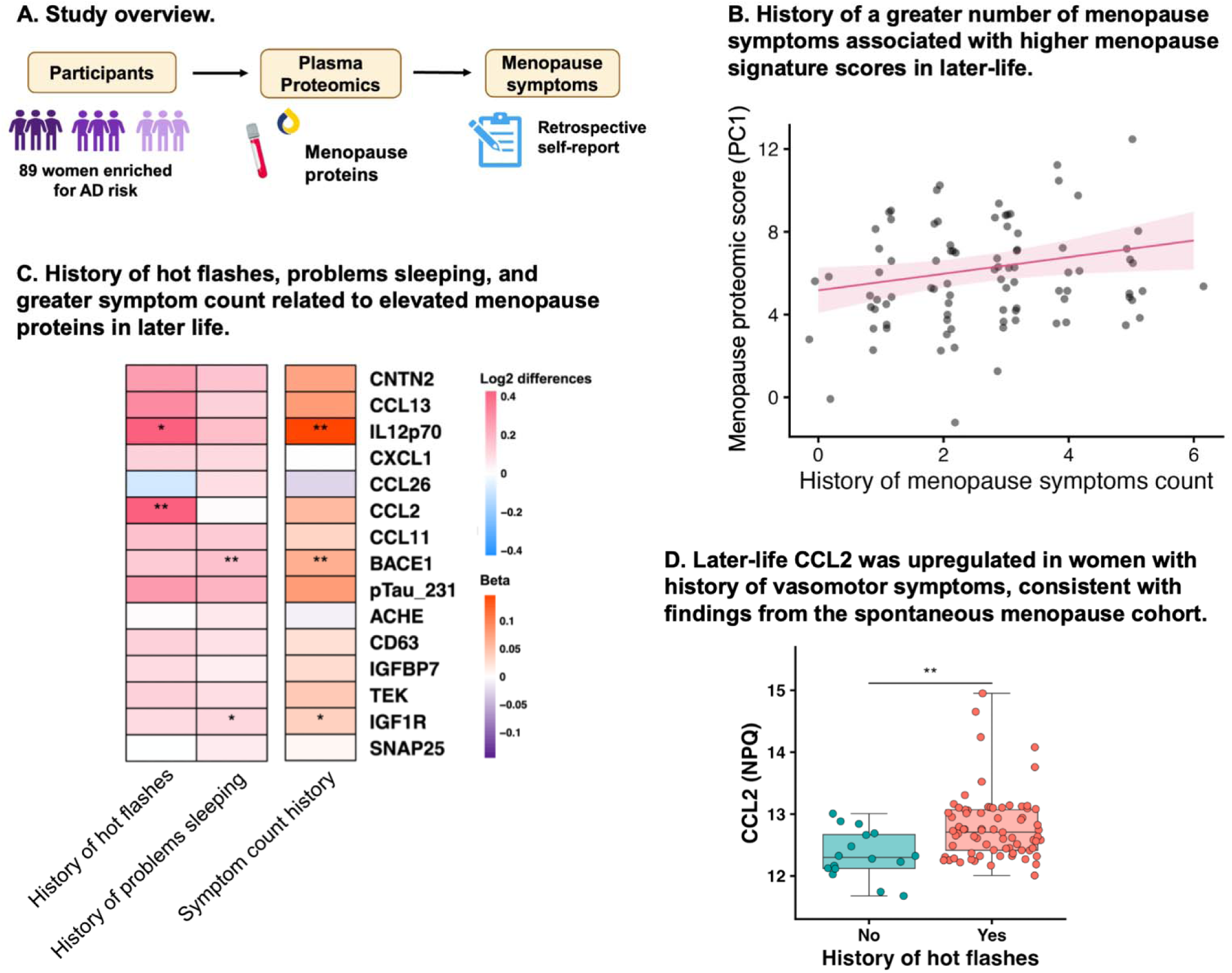
History of menopause symptoms relates to menopause protein levels years to decades later. **A.** In the aging cohort with menopause symptom data (WRAP), women self-reported their history of menopause symptoms, including hot flashes, night sweats, sexual dysfunction, problems sleeping, mood swings, and depression. Analyses evaluated associations of menopause symptom history with menopause scores and individual proteins, adjusted for age, history of hormone therapy, and bilateral oophorectomy. **B.** History of greater total menopause symptom count was associated with higher menopause proteomic scores in analyses adjusted for age, history of hormone therapy, and bilateral oophorectomy. **C.** Among symptoms that significantly associated with menopause scores (i.e., hot flashes, problems sleeping, and total symptom count), post hoc analyses tested associations with individual menopause proteins, adjusted for age, history of hormone therapy, and bilateral oophorectomy. History of hot flashes associated with increased IL12p70 and CCL2, problems sleeping with increased BACE1 and IGF1R, and symptom count with increased IL12p70, BACE1, and IGF1R. **D.** A boxplot visualization of CCL2 level by history of hot flashes showed higher levels in women with hot flashes vs. without. \**p*<.05, \*\**p*<.01, \*\*\**p*<.001.

#### Menopause proteomic changes related to cognitive aging and dementia risk

To test the relevance of proteomic shifts during menopause for later-life cognitive health, we computed the menopause proteomic score in four independent cohorts of older women (Tab.S10) with mean baseline age ranging from 60.7 to 72.1 years (Fig.6A). In longitudinal data with an average 5-year follow up from the Alzheimer’s Disease Neuroimaging Initiative (ADNI; *N*=673; 54.3% mild cognitive impairment [MCI]/dementia; 49.0% amyloid-positive) and UCSF Brain Aging Network for Cognitive Health (BrANCH; *N*=100; 1% MCI; 21.7% amyloid-positive), higher (i.e., more postmenopausal) baseline menopause proteomic scores associated with faster global cognitive decline (ADNI: β=-0.30, 95% CI=-0.42, -0.18, *p*<.001; BrANCH: β=-0.03, 95% CI=-0.05, -0.002, *p*=.03; Fig.6B-C). In cross-sectional data from WRAP (*N*=93; 15.0% MCI/dementia; 64.5% amyloid-positive), higher menopause proteomic scores associated with worse global performance (β=-0.27, 95% CI=-0.51, -0.04, *p*=.02; Fig.6D). Finally, in the UKB (*N*=11,059), we examined whether menopause proteomic scores associated with risk for incident dementia over a mean (*SD*) of 15.7 (2.84) years of follow-up. Higher menopause scores associated with greater risk for AD dementia (HR=1.15, 95% CI=1.01, 1.31, *p*=.04, *N*=219 events; Fig.6E), but not frontotemporal (HR=0.90, 95% CI=0.59, 1.36, *p*=.61, *N*=23 events), vascular (HR=1.15, 95% CI=0.91, 1.47, *p*=.25, *N*=65 events) or all-cause dementia (HR=1.08, 95% CI=0.98, 1.19, *p*=.13, *N*=387 events). Post hoc analyses showed that the individual menopause proteins associated with cognitive outcomes varied across cohorts, suggesting a broader coordinated biological relationship (versus individual proteins) (Fig.6F).

**Figure 6.**
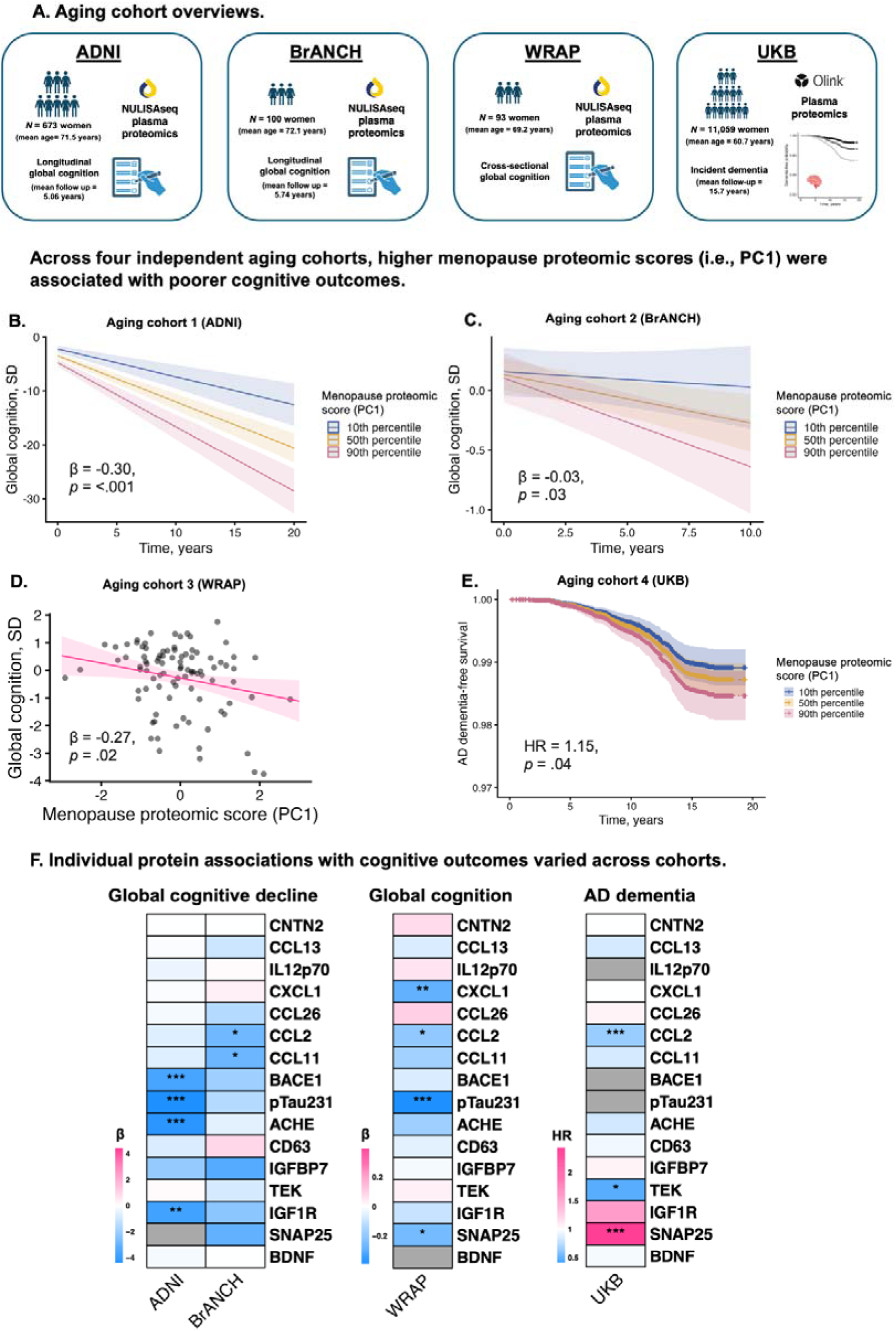
In later life, elevated menopause proteomic scores associated with poorer cognitive outcomes. **A.** Across four aging cohorts (i.e., the Alzheimer’s Disease Neuroimaging Initiative [ADNI; aging cohort 1], the Brain Aging Network for Cognitive Health [BrANCH; aging cohort 2], the Wisconsin Registry for Alzheimer’s Prevention [WRAP; aging cohort 3], and the UK Biobank [UKB; aging cohort 4]), we tested associations of the menopause proteomic score (i.e., PC1) with cognitive outcomes. **B-E.** In all cohorts, higher (i.e., more postmenopausal) menopause proteomic scores associated with poorer cognitive outcomes, including faster global cognitive decline in ADNI and BrANCH (**B-C**), lower global cognitive scores in WRAP (**D**), and greater risk for incident Alzheimer’s disease (AD) dementia in UKB (**E**). **F.** Individual menopause protein associations with cognitive outcomes varied across cohorts.

## Sensitivity analyses

Sensitivity analyses are described in the Extended Data and presented in Tables S14-16. In brief, findings were robust to (1) further restriction of the UCSB HAS sample by chronological age (i.e., 47–53; *N*=54; Fig.S6), (2) adjustment for cardiometabolic comorbidities, and in aging cohorts only: (3) exclusion of participants with bilateral oophorectomy, (4) stratification by history of hormone therapy, (5) exclusion of p-tau231 from the menopause proteomic score, and (6) exclusion of participants with MCI or dementia at baseline. Exploratory analyses testing effect modification by *APOE4* status showed largely null findings, with the exception that *APOE4* carriage significantly exacerbated the associations of the menopause proteomic score with cognitive decline in ADNI (Fig.S7).

## DISCUSSION

Applying attamolar-sensitivity, multiplex proteomics of brain aging in a rigorously staged cohort of midlife women, we identified molecular signals of menopause enriched for inflammatory, metabolic, synaptic, and AD biology. Menopause-related proteomic shifts tracked more strongly with hormones than with age. Across perimenopausal, early postmenopausal, and older women, vasomotor symptoms related to exaggerated increases in pro-inflammatory proteins. Top menopause proteomic shifts robustly replicated across analytic platforms in a large, independent cohort of age-matched pre/peri- and postmenopausal women, revealing wider menopause-associated upregulation of molecular pathways reflecting multisystem accelerated biological aging, and notably brain and oligodendrocyte aging. Further, in four independent cohorts of older postmenopausal women, menopause proteomics consistently associated with poorer cognitive outcomes, suggesting that menopause-related biological shifts may remain relevant years to decades post-menopause. Collectively, our findings highlight potential biology linking menopause to risk and resilience in cognitive aging.

The predominance of immune/inflammatory molecules associated with the menopause transition (e.g., CCL2, CCL11, TEK) alongside upregulation of pathways related to cytokine signalling and the complement cascade align with observed rises in systemic inflammation post-menopause.^2,23^ We also identify menopause-associated shifts in synaptic markers (ACHE, SNAP25, CNTN2), and are among the first to link menopause to blood markers of AD biology (p-tau231, BACE). These findings are consistent with animal models demonstrating widespread disruptions in synaptic plasticity, spine density, and cholinergic signalling^4,26,27^ and extend recent studies showing elevations in Aβ and tau-PET across menopause.^28–30^ Although all women with an ovary undergo menopause, most do not develop AD, and the clinical relevance of subthreshold elevations in p-tau231 and BACE1 in midlife is unclear. Overall, our findings align with previous observations of accelerated biological aging across menopause^31^ and underscore specific links to brain-relevant targets, overall brain aging, and oligodendrocyte age.

Menopause proteomic differences tracked more strongly with hormones than age, suggesting that the observed biological changes may be primarily driven by ovarian versus chronological aging. Many proteins were most strongly linked to FSH, including key targets implicated in AD pathobiology and synaptic processes (i.e., BACE1, ACHE, CNTN2).^32–34^ FSH receptors are abundantly expressed in AD-vulnerable brain regions, and preclinical models show that FSH can trigger development of AD pathology, neuroinflammation, and cognitive deficits.^35^ In contrast, lower estradiol was linked to increases in pro-inflammatory markers (CCL26, CCL2, TEK), aligning with the established role of estrogens in modulating immune responses.^36^

Vasomotor symptoms were associated with menopause proteomic scores, across both midlife and older women with histories of vasomotor symptoms. These associations were driven primarily by pro-inflammatory targets, with CCL2 (i.e., MCP-1) the most replicable. Vasomotor symptom severity has previously been associated with vascular inflammation^37,38^ (including CCL2^39^), cardiovascular risk, and AD brain changes.^20,21^ Vasomotor symptoms may signal less adaptive physiological responses to menopause that confer risk for adverse brain aging and other chronic diseases. It is unclear why only night sweats (not hot flashes) were associated with the menopause score in midlife women, and only hot flashes (not night sweats) were associated with the menopause score in the later-life cohort. Given their shared etiology,^10^ this inconsistency may relate to recall bias and imprecision introduced by self-report measures. There were also unexpected associations of vaginal dryness with lower menopause scores, the mechanistic significance of which is unclear.

Our study provides evidence that menopause-related proteomic changes are detectable and linked to cognitive trajectories in later life. Across four independent older adult cohorts, menopause proteomic scores associated with worse cognitive outcomes, including faster cognitive decline and greater risk of AD dementia. The effect size for AD dementia was relatively small, and the associations of individual proteins with cognitive outcomes varied across cohorts. These findings may reflect differences in cohort composition (e.g., age, cognitive status, amyloid positivity), as well as proteomic platform differences (NULISAseq/Olink). However, these results could also suggest that associations between menopause proteomic changes and cognitive outcomes are driven less by specific targets than by coordinated shifts that may generally reflect pro-aging biology.^40–42^

Strengths of this study include the unique midlife cohort with rigorously defined menopause stages to identify proteomic signals of menopause which largely replicated in a cross-platform, validation cohort of post-menopausal women age-matched to pre-peri-menopausal women. The age-matched validation provides compelling evidence that the observed proteomic changes were driven more strongly by endocrine than chronological aging. Three of the targets identified in the UCSB cohort did not show concordant elevations in the UKB sample (BDNF, CD63, CCL26), which may reflect proteomic platform differences, spurious findings, or coarse evaluation of menopause status in the UKB. Of note, while we used a composite score to summarize menopause-associated biological signals observed on ultrasensitive NULISAseq in our deeply-phenotyped cohort, the menopause proteomic score does not represent the full breadth of menopause-related biology detectable in circulation, as evidenced by the widespread proteomic shifts observed in the UKB Olink analyses. That is, NULISAseq CNS captures only ∼120 proteins involved in neurologic aging, while other high-throughput assays may allow expanded discovery. Furthermore, while serum and plasma appear to perform comparably on the NULISAseq CNS panel,^43^ matrix differences may influence protein measurements making cross-cohort comparison more challenging. Other limitations include the small spontaneous menopause discovery sample which only included cross-sectional protein and hormone data, the reliance on self-reported menopause symptoms, and that proteins in blood reflect multiple tissue sources. Longitudinal studies mapping within-person trajectories of hormones, multi-omics, symptoms, and brain health across menopause will be critical to confirm the observed menopause-associated proteomic changes and disentangle the complex interplay between endocrine, neurologic, and chronological aging.

We defined blood-based proteomic signals of menopause, enriched for inflammatory, metabolic, synaptic, AD biology, and accelerated biological aging. We demonstrated that inflammatory changes are especially pronounced in women with vasomotor symptoms, highlighting CCL2 as a signal of this physiologic susceptibility that may persist into later life. We further illustrated that menopause biology shows continued relevance to cognitive aging across four independent cohorts of older women. These findings shine new light on the biological processes connecting menopause to dementia risk, affirming the importance of menopause as a critical window for our foundational understanding of brain aging.

## METHODS

### Spontaneous menopause cohort participants

We recruited approximately equal numbers of pre-, peri-, and postmenopausal cisgender women (*N*=80) plus age-matched cisgender men (*N*=36) to participate in the UCSB Healthy Aging Study (HAS). Menopause was staged following STRAW+10 criteria.^24^ Participants were aged 35–60, not currently taking menopausal hormone therapy, and did not have any major medical conditions. Participants who were not taking hormonal contraceptives were prioritized for recruitment, resulting in a sample that was largely free of exogenous hormones (*N*=1 on oral contraceptives). Female participants were required to have an intact uterus and no history of bilateral oophorectomy. Female participants self-reported how often they experienced commonly endorsed menopause symptoms^44,45^ including hot flashes, night sweats, vaginal dryness, and irritability in the last two weeks, on a scale of “not at all”, “1-5 days”, “6-8 days”, “9-13 days”, or “every day”. Due to the small sample size, each individual symptom was dichotomized as present versus absent.

### Endocrine assays

Serum from a venous blood draw was analyzed for gonadotropin and sex steroid hormones: FSH, 17β-estradiol, progesterone, sex-hormone binding globulin (SHBG), dehydroepiandrosterone sulfate (DHEAS), and testosterone (Extended Data).

### Blood proteomics

Serum (menopause cohort) or plasma (aging cohorts) was analyzed on the NULISAseq CNS panel to measure 127 proteins in UCSB HAS, 131 in ADNI, 132 in BrANCH, and 122 in WRAP using the automated Alamar Argo HT workflow (Extended Data).^25,46,47^

### Cross-cohort, cross-platform validation in UK Biobank

To evaluate whether spontaneous menopause proteomic shifts were robust to cohort and proteomic platform, we compared spontaneous menopause protein effect sizes detected in the midlife menopause cohort with NULISAseq proteomics to menopause protein effect sizes detected in a large cohort of 2,814 women from UK Biobank with cross-sectional proximity extension assay Olink proteomics (Extended Data).^48^ Pre/peri-menopausal (*N*=1,407) and postmenopausal (*N*=1,407) women were propensity matched on age within a range of 45-60 years, comparable to the UCSB cohort.

### Aging cohorts

To evaluate the relevance of menopause-related biological changes to later-life cognitive outcomes, we used proteomics data from women in four independent aging cohorts (Extended Data). These included the Alzheimer’s Disease Neuroimaging Initiative (ADNI; *N*=673), the UCSF Brain Aging Network for Cognitive Health (BrANCH; *N*=100), the Wisconsin Registry for Alzheimer’s Prevention (WRAP; *N*=93), and the UK Biobank (UKB; *N*=11,059). ADNI, BrANCH, and WRAP had plasma NULISAseq proteomics, and UKB had plasma Olink proteomics. We examined longitudinal global cognitive decline in ADNI and BrANCH, and cross-sectional global cognition in WRAP due to limited follow-up. In the UKB, we examined incident dementia. Studies were approved by research ethics boards and all participants provided informed consent.

## Analyses

### Spontaneous menopause cohort

Age-adjusted ANCOVA compared levels of 126 NULISAseq proteins by menopause stage (pre/peri/post). Menopause proteins were identified as those that differed between pre- and postmenopausal women per nominal statistical significance (unadjusted *p*<.05). All further analyses focused exclusively on these proteins. We performed a PCA on the identified menopause proteins in all midlife women and extracted the first component (PC1) as a “menopause proteomic score.” To limit comparisons, the menopause score was used as the primary measure for all further analyses, with effects of individual proteins probed in post hoc analyses. Separate age-adjusted regressions tested the associations of menopause stage and sex with the menopause score.

While menopause definitionally involves endocrine shifts, different hormones may relate to unique aspects of menopause biology. To probe this, age-adjusted regressions tested associations between sex hormones and the menopause proteomic score in all midlife women. Among peri- and post-menopausal women, separate regressions tested associations of menopause symptoms with the menopause score, adjusting for age and menopause stage.

### Cross-cohort, cross-platform validation

In the age-matched sample of pre/peri- and postmenopausal women in the UKB, age-adjusted regression models tested associations between menopause status (post vs. pre/peri) and 2,923 Olink proteins, with FDR correction. For the menopause proteins identified in UCSB (NULISAseq) that were measured on Olink, we examined whether these proteins also differed between pre/peri- and postmenopausal women in the UKB, including estimated proteomic effect sizes between cohorts. We performed GO and pathway enrichment and tested associations of menopause stage with proteomic clocks of organ and cellular aging^49,50^ (Extended Data).

### Aging cohorts

We extracted loadings and summary statistics for the menopause score (PC1) calculated in the spontaneous menopause cohort to compute an equivalent score in each aging cohort (Extended Data). In WRAP, regressions tested associations of menopause symptom history with the menopause score, adjusting for age, history of hormone therapy, and bilateral oophorectomy.

To examine the relevance of the menopause score to cognitive aging outcomes, in ADNI and BrANCH, mixed effects models tested associations of the interaction between the score and time on global cognition, adjusting for baseline age and education. In WRAP, a linear regression tested the association of menopause score on global cognition, adjusting for age and education. In the UKB, Cox proportional-hazards models tested associations of the menopause score with risk for incident dementia (AD, frontotemporal, vascular, all-cause), adjusting for age.

## Data Availability

All data produced in this work are available upon reasonable request to the included studies.

## Funding/Acknowledgments

This research has been conducted using the UK Biobank Resource under Application Number 1045037. This work was supported by the Wellcome Leap CARE program (MPI: KBC, RS, EGJ), NIH-NIA grants R01AG032289 (PI: JHK), R01AG048234 (PI: JHK), R01 AG063843 (EGJ); RF1AG096165 (KC); RF1AG096477 (EGJ); the Noyce Trust (NM, EGJ); the Ann S. Bowers Women’s Brain Health Initiative (NM, KC, EGJ); UCSF ADRC P30AG062422 (PI: GDR), R01AG072475 (PI: KBC), and K23AG090757 (PI: RS). WRAP is supported by AG027161 (PI: SCJ). Our work is also supported by a grant from the Larry L. Hillblom Foundation (2024-A-001-CTR; PI: KBC) and previously by grant (2018-A-006-NET; PI: JHK), New Vision Research (CCAD 2024-001-1; PI: RS), an award from the American Academy of Neurology (PI: RS), funding from the Alzheimer Society of Canada Research Program (MWA, JSR), CIHR (173253, 438475, Canada Graduate Scholarships program; JSR, MWA), the Alzheimer’s Association (JSR); NINDS UE5NS070680 (PI: SAJ, Recipient: MC), the Noyce Trust (PI: NM), and NIH/NIA K23AG084883 (EP). This work was supported in part by NIA Intramural Research Program (K.A.W.). The contributions of the NIH author(s) are considered Works of the United States Government. The findings and conclusions presented in this paper are those of the author(s) and do not necessarily reflect the views of the NIH or the U.S. Department of Health and Human Services. Work on *The effects of pharmacological and spontaneous menopause on Alzheimer’s risk in women* is supported by Wellcome Leap as part of the CARE Program. Plots were created in part using Biorender.

## References

1. Levine ME, Lu AT, Chen BH, Hernandez DG, Singleton AB, Ferrucci L, et al. Menopause accelerates biological aging. Proc Natl Acad Sci U S A. 2016 Aug 16;113(33):9327–32. doi:10.1073/pnas.1604558113 PubMed PMID: 27457926; PubMed Central PMCID: PMC4995944.

2. McCarthy M, Raval AP. The peri-menopause in a woman’s life: a systemic inflammatory phase that enables later neurodegenerative disease. J Neuroinflammation. 2020 Oct 23;17(1):317. doi:10.1186/s12974-020-01998-9 PubMed PMID: 33097048; PubMed Central PMCID: PMC7585188.

3. El Khoudary SR, Aggarwal B, Beckie TM, Hodis HN, Johnson AE, Langer RD, et al. Menopause Transition and Cardiovascular Disease Risk: Implications for Timing of Early Prevention: A Scientific Statement From the American Heart Association. Circulation. 2020 Dec 22;142(25):e506–32. doi:10.1161/CIR.0000000000000912 PubMed PMID: 33251828.

4. Hara Y, Waters EM, McEwen BS, Morrison JH. Estrogen Effects on Cognitive and Synaptic Health Over the Lifecourse. Physiol Rev. 2015 Jul;95(3):785–807. doi:10.1152/physrev.00036.2014 PubMed PMID: 26109339; PubMed Central PMCID: PMC4491541.

5. Wood Alexander M, Honer WG, Saloner R, Galea LAM, Bennett DA, Rabin JS, et al. The interplay between age at menopause and synaptic integrity on Alzheimer’s disease risk in women. Sci Adv. 2025 Mar 7;11(10):eadt0757. doi:10.1126/sciadv.adt0757 PubMed PMID: 40043118; PubMed Central PMCID: PMC11881898.

6. Coughlan GT, Betthauser TJ, Boyle R, Koscik RL, Klinger HM, Chibnik LB, et al. Association of Age at Menopause and Hormone Therapy Use With Tau and β-Amyloid Positron Emission Tomography. JAMA Neurol. 2023 Apr 3;e230455. doi:10.1001/jamaneurol.2023.0455 PubMed PMID: 37010830; PubMed Central PMCID: PMC10071399.

7. Gong J, Harris K, Peters SAE, Woodward M. Reproductive factors and the risk of incident dementia: A cohort study of UK Biobank participants. Brayne C, editor. PLoS Med. 2022 Apr 5;19(4):e1003955. doi:10.1371/journal.pmed.1003955

8. Wood Alexander M, Wu CY, Coughlan GT, Puri T, Buckley RF, Palta P, et al. Associations Between Age at Menopause, Vascular Risk, and 3-Year Cognitive Change in the Canadian Longitudinal Study on Aging. Neurology. 2024 May 14;102(9):e209298. doi:10.1212/WNL.0000000000209298

9. Wood Alexander M, Fischer DL, VandeVrede L, Nichols E, Yu L, Pike JR, et al. Sex Differences in Associations of Lewy Body Disease with Alzheimer’s Disease and Cognitive Decline. Ann Neurol. 2025 Jul 11. doi:10.1002/ana.27308 PubMed PMID: 40646704; PubMed Central PMCID: PMC12341763.

10. Thurston RC, Thomas HN, Castle AJ, Gibson CJ. Menopause as a biological and psychological transition. Nat Rev Psychol. 2025 Jun 20;4(8):530–43. doi:10.1038/s44159-025-00463-9

11. Maki PM, Thurston RC. Menopause and Brain Health: Hormonal Changes Are Only Part of the Story. Front Neurol. 2020;11:562275. doi:10.3389/fneur.2020.562275 PubMed PMID: 33071945; PubMed Central PMCID: PMC7538803.

12. Gurvich C, Spector A, Hickey M. Advances in understanding of cognitive symptoms during menopause. The Lancet Obstetrics, Gynaecology, & Women’s Health. 2026 Apr;2(4):e335–45. doi:10.1016/S3050-5038(26)00043-9

13. Downs JL, Wise PM. The role of the brain in female reproductive aging. Mol Cell Endocrinol. 2009 Feb 5;299(1):32–8. doi:10.1016/j.mce.2008.11.012 PubMed PMID: 19063938; PubMed Central PMCID: PMC2692385.

14. Bacon ER, Mishra A, Wang Y, Desai MK, Yin F, Brinton RD. Neuroendocrine aging precedes perimenopause and is regulated by DNA methylation. Neurobiol Aging. 2019 Feb;74:213–24. doi:10.1016/j.neurobiolaging.2018.09.029 PubMed PMID: 30497015; PubMed Central PMCID: PMC7117064.

15. Jack CR, Bennett DA, Blennow K, Carrillo MC, Dunn B, Haeberlein SB, et al. NIA-AA Research Framework: Toward a biological definition of Alzheimer’s disease. Alzheimer’s & Dementia. 2018 Apr;14(4):535–62. doi:10.1016/j.jalz.2018.02.018

16. Sperling RA, Aisen PS, Beckett LA, Bennett DA, Craft S, Fagan AM, et al. Toward defining the preclinical stages of Alzheimer’s disease: Recommendations from the National Institute on Aging-Alzheimer’s Association workgroups on diagnostic guidelines for Alzheimer’s disease. Alzheimer’s & Dementia. 2011 May;7(3):280– 92. doi:10.1016/j.jalz.2011.03.003

17. Gold EB. The Timing of the Age at Which Natural Menopause Occurs. Obstet Gynecol Clin North Am. 2011 Sep;38(3):425–40. doi:10.1016/j.ogc.2011.05.002 PubMed PMID: 21961711; PubMed Central PMCID: PMC3285482.

18. Bove R, Secor E, Chibnik LB, Barnes LL, Schneider JA, Bennett DA, et al. Age at surgical menopause influences cognitive decline and Alzheimer pathology in older women. Neurology. 2014 Jan 21;82(3):222–9. doi:10.1212/WNL.0000000000000033 PubMed PMID: 24336141; PubMed Central PMCID: PMC3902759.

19. Maki PM, Drogos LL, Rubin LH, Banuvar S, Shulman LP, Geller SE. Objective hot flashes are negatively related to verbal memory performance in midlife women. Menopause. 2008 Sep;15(5):848–56. doi:10.1097/gme.0b013e31816d815e

20. Thurston RC, Wu M, Chang YF, Aizenstein HJ, Derby CA, Barinas-Mitchell EA, et al. Menopausal Vasomotor Symptoms and White Matter Hyperintensities in Midlife Women. Neurology. 2023 Jan 10;100(2):e133–41. doi:10.1212/WNL.0000000000201401 PubMed PMID: 36224031; PubMed Central PMCID: PMC9841446.

21. Thurston RC, Maki P, Chang Y, Wu M, Aizenstein HJ, Derby CA, et al. Menopausal Vasomotor Symptoms and Plasma Alzheimer’s Disease Biomarkers. Am J Obstet Gynecol. 2024 Mar;230(3):342.e1-342.e8. doi:10.1016/j.ajog.2023.11.002 PubMed PMID: 37939982; PubMed Central PMCID: PMC10939914.

22. Appiah D, Schreiner PJ, Pankow JS, Brock G, Tang W, Norby FL, et al. Long-term changes in plasma proteomic profiles in premenopausal and postmenopausal Black and White women: the Atherosclerosis Risk in Communities study. Menopause. 2022 Oct 1;29(10):1150–60. doi:10.1097/GME.0000000000002031 PubMed PMID: 35969495; PubMed Central PMCID: PMC9509415.

23. De Maeyer RPH, Sikora J, Bracken OV, Shih B, Lloyd AF, Peckham H, et al. Age-Associated Inflammatory Monocytes Are Increased in Menopausal Females and Reversed by Hormone Replacement Therapy. Aging Cell. 2025 Nov;24(11):e70249. doi:10.1111/acel.70249

24. Harlow SD, Gass M, Hall JE, Lobo R, Maki P, Rebar RW, et al. Executive summary of the Stages of Reproductive Aging Workshop + 10: addressing the unfinished agenda of staging reproductive aging. Menopause. 2012 Apr;19(4):387–95. doi:10.1097/gme.0b013e31824d8f40 PubMed PMID: 22343510; PubMed Central PMCID: PMC3340903.

25. Zeng X, Lafferty TK, Sehrawat A, Chen Y, Ferreira PCL, Bellaver B, et al. Multi-analyte proteomic analysis identifies blood-based neuroinflammation, cerebrovascular and synaptic biomarkers in preclinical Alzheimer’s disease. Mol Neurodegeneration. 2024 Oct 10;19(1):68. doi:10.1186/s13024-024-00753-5

26. Salathe SF, Franczak E, Busick Z, Boakye FB, Allen J, Lutkewitte A, et al. Loss of ovarian function and estrogen therapy remodel the brain’s synaptic and metabolic proteome [Internet]. Neuroscience; 2025 [cited 2025 Nov 20]. Available from: http://biorxiv.org/lookup/doi/10.1101/2025.10.20.683488doi:10.1101/2025.10.20.683488

27. Russell JK, Jones CK, Newhouse PA. The Role of Estrogen in Brain and Cognitive Aging. Neurotherapeutics. 2019 Jul 15;16(3):649–65. doi:10.1007/s13311-019-00766-9

28. Buckley RF, O’Donnell A, McGrath ER, Jacobs HIL, Lois C, Satizabal CL, et al. Menopause Status Moderates Sex Differences in Tau Burden: A Framingham PET Study. Annals of Neurology. 2022;92(1):11–22. doi:10.1002/ana.26382

29. Rahman A, Schelbaum E, Hoffman K, Diaz I, Hristov H, Andrews R, et al. Sex-driven modifiers of Alzheimer risk. Neurology. 2020 Jul 14;95(2):e166–78. doi:10.1212/WNL.0000000000009781 PubMed PMID: 32580974; PubMed Central PMCID: PMC7455325.

30. Mosconi L, Berti V, Quinn C, McHugh P, Petrongolo G, Varsavsky I, et al. Sex differences in Alzheimer risk. Neurology. 2017 Sep 26;89(13):1382–90. doi:10.1212/WNL.0000000000004425 PubMed PMID: 28855400; PubMed Central PMCID: PMC5652968.

31. Xiang Y, Meng Q, Huang Z, Zhang N, Zhang Y, Ding X, et al. Menopausal status, transition, and age at menopause with accelerated biological aging across multiple organ systems: findings from two cohort studies. BMC Med. 2025 Aug 6;23(1):461. doi:10.1186/s12916-025-04223-7 PubMed PMID: 40770753; PubMed Central PMCID: PMC12330081.

32. Cole SL, Vassar R. The Alzheimer’s disease Beta-secretase enzyme, BACE1. Mol Neurodegeneration. 2007;2(1):22. doi:10.1186/1750-1326-2-22

33. Talesa VN. Acetylcholinesterase in Alzheimer’s disease. Mechanisms of Ageing and Development. 2001 Nov;122(16):1961–9. doi:10.1016/S0047-6374(01)00309-8

34. Chatterjee M, Del Campo M, Morrema THJ, De Waal M, Van Der Flier WM, Hoozemans JJM, et al. Contactin-2, a synaptic and axonal protein, is reduced in cerebrospinal fluid and brain tissue in Alzheimer’s disease. Alz Res Therapy. 2018 Dec;10(1):52. doi:10.1186/s13195-018-0383-x

35. Xiong J, Kang SS, Wang Z, Liu X, Kuo TC, Korkmaz F, et al. FSH blockade improves cognition in mice with Alzheimer’s disease. Nature. 2022 Mar 17;603(7901):470–6. doi:10.1038/s41586-022-04463-0

36. Straub RH. The Complex Role of Estrogens in Inflammation. Endocrine Reviews. 2007 Aug 1;28(5):521–74. doi:10.1210/er.2007-0001

37. Thurston RC, El Khoudary SR, Sutton-Tyrrell K, Crandall CJ, Gold E, Sternfeld B, et al. Are vasomotor symptoms associated with alterations in hemostatic and inflammatory markers? Findings from the Study of Women’s Health Across the Nation. Menopause. 2011 Oct;18(10):1044–51. doi:10.1097/gme.0b013e31821f5d39 PubMed PMID: 21926929; PubMed Central PMCID: PMC3183159.

38. Bechlioulis A, Naka KK, Kalantaridou SN, Kaponis A, Papanikolaou O, Vezyraki P, et al. Increased Vascular Inflammation in Early Menopausal Women Is Associated with Hot Flush Severity. The Journal of Clinical Endocrinology & Metabolism. 2012 May;97(5):E760–4. doi:10.1210/jc.2011-3151

39. Thurston RC, Chang Y, Mancuso P, Matthews KA. Adipokines, adiposity, and vasomotor symptoms during the menopause transition: findings from the Study of Women’s Health Across the Nation. Fertil Steril. 2013 Sep;100(3):793–800. doi:10.1016/j.fertnstert.2013.05.005 PubMed PMID: 23755948; PubMed Central PMCID: PMC3759568.

40. Nazarinia D, Behzadifard M, Gholampour J, Karimi R, Gholampour M. Eotaxin-1 (CCL11) in neuroinflammatory disorders and possible role in COVID-19 neurologic complications. Acta Neurol Belg. 2022 Aug;122(4):865–9. doi:10.1007/s13760-022-01984-3 PubMed PMID: 35690992; PubMed Central PMCID: PMC9188656.

41. Bettcher BM, Fitch R, Wynn MJ, Lalli MA, Elofson J, Jastrzab L, et al. MCP-1 and eotaxin-1 selectively and negatively associate with memory in MCI and Alzheimer’s disease dementia phenotypes. Alz & Dem Diag Ass & Dis Mo. 2016 Jan;3(1):91–7. doi:10.1016/j.dadm.2016.05.004

42. Orenduff MC, Pieper CF, Allott EH, Coleman MF, Jung SY, Vitolins MZ, et al. Plasma Insulin-like Growth Factor-Binding Protein-7 Is Positively Associated with Age, Obesity, Mortality, and Cancer in Postmenopausal Women. Cancer Epidemiol Biomarkers Prev. 2025 Jun 3;34(6):922–32. doi:10.1158/1055-9965.EPI-24-1644 PubMed PMID: 40152978; PubMed Central PMCID: PMC12724625.

43. Farinas MF, Chen Y, Zeng X, Nafash MN, Cohen AD, Lopez OL, et al. Evaluation of serum as an alternative matrix to plasma for NULISA based proteomic blood biomarker measurements. Sci Rep. 2026 Apr 18. doi:10.1038/s41598-026-46409-w PubMed PMID: 42000813.

44. Hedges MS, Hewings-Martin Y, Karam J, Castaneda R, Cunningham AC, Xu Y, et al. Global perspectives on perimenopause: a digital survey of knowledge and symptoms using the Flo application. Menopause. 2026 Jan 28. doi:10.1097/GME.0000000000002730 PubMed PMID: 41603602.

45. Fang Y, Liu F, Zhang X, Chen L, Liu Y, Yang L, et al. Mapping global prevalence of menopausal symptoms among middle-aged women: a systematic review and meta-analysis. BMC Public Health. 2024 Jul 2;24(1):1767. doi:10.1186/s12889-024-19280-5 PubMed PMID: 38956480; PubMed Central PMCID: PMC11220992.

46. Rea Reyes RE, Wilson RE, Langhough RE, Studer RL, Jonaitis EM, Oomens JE, et al. Targeted proteomic biomarker profiling using NULISA in a cohort enriched with risk for Alzheimer’s disease and related dementias. Alzheimers Dement. 2025 May;21(5):e70166. doi:10.1002/alz.70166 PubMed PMID: 40318118; PubMed Central PMCID: PMC12046973.

47. Feng W, Beer JC, Hao Q, Ariyapala IS, Sahajan A, Komarov A, et al. NULISA: a proteomic liquid biopsy platform with attomolar sensitivity and high multiplexing. Nat Commun. 2023 Nov 9;14(1):7238. doi:10.1038/s41467-023-42834-x

48. Sun BB, Chiou J, Traylor M, Benner C, Hsu YH, Richardson TG, et al. Plasma proteomic associations with genetics and health in the UK Biobank. Nature. 2023 Oct;622(7982):329–38. doi:10.1038/s41586-023-06592-6 PubMed PMID: 37794186; PubMed Central PMCID: PMC10567551.

49. Oh HSH, Le Guen Y, Rappoport N, Urey DY, Farinas A, Rutledge J, et al. Plasma proteomics links brain and immune system aging with healthspan and longevity. Nat Med. 2025 Aug;31(8):2703–11. doi:10.1038/s41591-025-03798-1

50. Ding DY, Bot VA, Chen KL, Groves JW, Pálovics R, Masuda D, et al. Plasma proteomic signatures of cellular aging predict human disease. Nat Med. 2026 Jun 15. doi:10.1038/s41591-026-04446-y

