## Extended data for "Blood proteomics of menopause map to brain aging and dementia risk"

**Extended methods**

**Serum hormone quantification in UCSB HAS (menopause cohort):**

All participants fasted for 8 hours before a morning venous blood draw. Women who were still menstruating were scheduled in the early follicular phase (day 3–5) of their menstrual cycle, pursuant to subject report.

Serum was analyzed for six gonadotropin and sex steroid hormones: follicle-stimulating hormone (FSH), 17β-estradiol, progesterone, sex-hormone binding globulin (SHBG), dehydroepiandrosterone sulfate (DHEAS), and testosterone at the Brigham and Women’s Hospital Research Assay Core. 17β-estradiol, progesterone, and testosterone were determined by liquid chromatography mass-spectrometry, and FSH, SHBG, and DHEAS by chemiluminescent immunoassay (Beckman Coulter). Assay sensitivities, dynamic range, and intra-assay coefficients of variation (respectively) were as follows: estradiol, 1 pg/mL, 1–500 pg/mL, <5% relative standard deviation (RSD); progesterone, 0.05 ng/mL, 0.05–10 ng/mL, 9.33% RSD; testosterone, 1.0 ng/dL, 1–2000 ng/dL, <4% RSD; FSH, 0.2 mIU/mL, 0.2–200 mIU/mL, 3.1-4.3%; SHBG, 0.33 nmol/L, 0.33-200 nmol/L, 4.5–4.8%; DHEAS, 2 ug/dL, 2–1000 ug/dL, 1.6–8.3%.

All 6 hormones were measured in women, and the latter three were measured in men. Hormone levels were log-transformed and expressed in standard deviations to aid interpretability.

**NULISA blood proteomics:**

Serum/plasma was analyzed on the NULISAseq CNS panel to measure 127 proteins in UCSB HAS, 131 in ADNI, 132 in BrANCH, and 122 in WRAP. The automated Alamar Argo HT workflow generated immunocomplexes tagged with both sample-specific and target-specific DNA barcodes. Samples were pooled into a single library for next-generation DNA sequencing. Samples were randomized (non-stratified) across plates and run in singlet, per previously established procedures for NULISA assays.^1,2^ Batch (plate) effects were mitigated using the standard NULISA Protein Quantification (NPQ) protocols and algorithm (as previously described),^1^ including hierarchical normalization that consists of internal spike-in controls for well-level normalization, pooled inter-plate controls to harmonize protein measurements across plates, and negative controls to estimate background and limits of detection. Protein concentrations are reported in NPQ units, which are computed by normalizing the relative concentration of each analyte using internal and inter-plate controls and log_2_-transforming the normalized values. Plate- and sample-level QC metrics (e.g., coefficient of variation (CV), detectability) were used to assess assay performance. The limit of detectability (LOD) is determined as the mean plus three times the standard deviation of the exponentiated (i.e., anti-log-transformed) normalized counts for the negative control samples on the plate, which are run in quadruplicates. Detectability is then calculated as the percentage of samples in which the quantification is above the LOD for that target*.*

In all cohorts, QC metrics were in acceptable ranges, including for both intra-plate internal control CV (UCSB HAS: 15.0%; BrANCH: 7.3%; ADNI: 5.9%; WRAP: 0.01-7.5%; all within maximum threshold of 25%) and inter-plate control CV (UCSB HAS: 4.0%; BrANCH: 8.6%; ADNI: 11.8%; WRAP: 0.29-5.6%; all within maximum threshold of 15%). In the UCSB HAS, 8 proteins (i.e., UCHL1, SNCB, PTDP43409, PTN, pTau217, OligoSNCA, IL1B, and NRGN) had median detectability of <50%, so were removed from analyses following previous work with the NULISA panel.^3^ In all other NULISA cohorts (i.e., BrANCH, ADNI), our analyses were restricted to the 16 menopause proteins identified in the UCSB HAS. All 16 of these menopause proteins were detectable (≥50%) in BrANCH and WRAP and all except SNAP25 were detectable (≥50%) in ADNI. SNAP25 was removed from analyses in ADNI. Given that APOE4 protein expression proxies *APOE4* genotype (bimodal distribution), we excluded APOE4 from proteomics analyses. In the UCSB HAS, APOE4 levels were used to infer *APOE4* allele carriage (i.e., APOE4 ≥10 NPQ categorized as carrier, APOE4 <10 NPQ categorized as non-carrier; Fig.S8). A full list of proteins in each cohort is provided in Table S17.

To reduce the influence of extreme values, all protein concentrations were winsorized at the three-interquartile limit. There were no missing values on any examined proteins in the UCSB HAS cohort. No imputation was used for NULISAseq data, and analyses in other NULISA cohorts (i.e., BrANCH, ADNI, WRAP) were restricted to participants with non-missing values on all menopause proteins. Given all analyses examined proteomic outcomes within cohorts separately, we did not normalize protein levels between cohorts.

**Cross-cohort, cross-platform menopause validation in UK Biobank:**

The UK Biobank is a population-based prospective cohort study with multi-omics and clinical data on approximately 500,000 participants aged 40-69 years. The UKB-PPP consortium generated proximity extension assay (PEA) Olink plasma proteomics data on a subset participants’ baseline samples, resulting in up to 2,923 protein measurements from 52,995 participants in the post-UKB-PPP quality control dataset. Details of proteomics data generation, processing, and quality control have been previously described^4^ and are available at <https://biobank.ndph.ox.ac.uk/ukb/ukb/docs/PPP_Phase_1_QC_dataset_companion_doc.pdf> and <https://biobank.ndph.ox.ac.uk/crystal/ukb/docs/Olink-3072_B0-B6_Analysis-Report.pdf>. In brief, samples were aliquoted in duplicate using a pseudo-randomisation algorithm to maximise variation of phenotypes on each generated sample plate, balancing factors such as age, sex, day of the week of blood collection, self-reported ethnicity, and deprivation index.^4^ QC metrics were all in acceptable ranges for Olink proteomics, including median intra-plate CVs were <8%, median inter-plate CVs <11%, median inter-instrument CVs<11%, and 73.2% of proteins had less than 30% of measurements falling below the LODs.^4^ Normalized Protein Expression (NPX) values were calculated per standard, previously described algorithms that include normalizing for both plate-to-plate and batch-to-batch variation. To maintain consistency with other published UKB proteomics papers,^4,5^ we used the standard, post-quality control UKB Olink dataset consisting of 2,923 unique proteins, including 13 menopause proteins (p-tau231, BACE1, and IL12p70 were not measured in UKB).

*N*=17,487 female participants reported being postmenopausal at baseline (answered “yes” to "Have you had your menopause (periods stopped)?"), and *N*=6,542 female participants reported not being postmenopausal at baseline (answered “no”; *N*=3,272 with hysterectomy and *N*=1,231 not sure/prefer not to answer excluded). Of the resultant sample of *N*=24,029 pre/peri- and postmenopausal women, we further restricted to participants who reported no history of bilateral oophorectomy (*N*=22,809; *N*=1,220 excluded), no current use of menopausal hormone therapy (*N*=21,910; *N*=899 excluded); and were aged 45-60 at the time of the Olink blood draw to more closely mirror the UCSB HAS sample (*N*=10,926 [*N*=3,789 pre/peri-menopausal, *N*=7,137 postmenopausal]; *N*=10,984 excluded).

To mitigate the confound between age and menopause status, we created age-matched samples of pre- and postmenopausal women (R “MatchIt” package). Pre/peri- and postmenopausal women were matched with a 1:1 ratio using nearest-neighbour matching without replacement and a 0.05 *SD* caliper. This resulted in a sample of N=1,407 premenopausal women and N=1,407 postmenopausal women who were nearly exactly matched on age (Figure 4A, Table S7) for validation analyses.

**Aging cohorts**

***Alzheimer’s Disease Neuroimaging Initiative (ADNI; aging cohort 1):*** ADNI is an international multi-site observational study that follows older adults across the cognitive spectrum (i.e., cognitively normal, mild cognitive impairment [MCI], and AD).^6^ No data were collected on menopause history or symptoms. Participants underwent repeated cognitive assessments and blood collection every 6-12 months. To quantify global cognition, we examined the ADNI modified Preclinical Alzheimer’s Cognitive Composite (mPACC) score, which was computed by averaging sample-based z-scores on the Alzheimer Disease Assessment Scale—Cognitive Subscale Delayed Word Recall, Logical Memory Delayed Recall, the Mini Mental State Examination (MMSE), and (log-transformed) Trail-Making Test B Time to Completion.^7^ For a subset of ADNI participants (*N*=1,248), baseline plasma was analyzed for 131 proteins on the NULISAseq CNS platform. Our analytic sample included *N*=673 female participants who had NULISAseq data and cognitive test data available. To characterize the sample, amyloid positivity was determined using the derived ADNI variable “AMYSTAT”, which uses validated, tracer-specific quantitative thresholds to classify participants’ amyloid status as “elevated” or “not elevated’ from harmonized Aβ positron emission tomography (PET) data.^8,9^

***UCSF Brain Aging Network for Cognitive Health (BrANCH; aging cohort 2):*** BrANCH is a study of community-dwelling, functionally intact participants who undergo annual neuropsychological testing and blood draws. Female participants self-reported hormone therapy use and oophorectomy. A composite score of global cognition was computed by averaging sample-based z-scores for tests of memory (California Verbal Learning Test II immediate and delayed recall), executive function (Stroop interference, modified Trail Making Test, phonemic fluency, and Digit Span Backwards), and processing speed (six computerized visually based tests of Length Judgment, Visual Search, Distance Judgment, Abstract Matching 1, Abstract Matching 2, and Shape Judgment), described previously.^10^ Baseline plasma was also analyzed for 132 proteins on the NULISAseq CNS platform. Our analytic sample included *N*=100 female participants with NULISAseq who had cognitive data available. Baseline plasma p-tau181/Aβ42 was assayed on Simoa (Quanterix). To characterize the sample, amyloid positivity was determined using a sample-specific, Plasma p-tau181/Aβ42 was assayed on Simoa (Quanterix) across three batches in the broader BrANCH cohort (1,266 samples across *N*=664 participants). Batch effects were quantified via linear regression on samples shared across batches, with model estimates subsequently applied to all samples to align cross-batch values on the same scale. Amyloid positivity was determined using a sample-specific, batch-harmonized p-tau181/Aβ42 threshold that optimized classification of visually read Aβ-PET 18F-AV45 positivity in the broader BrANCH cohort (AUC=0.88).

***Wisconsin Registry for Alzheimer’s Prevention (WRAP; aging cohort 3):*** WRAP is an observational study enriched for participants with a parental history of AD. Participants completed clinical/cognitive testing and blood collection approximately every two years. Female participants self-reported histories of hormone therapy use, oophorectomy, and menopause symptoms. Menopause symptoms reported included hot flashes, night sweats, sexual dysfunction, problems sleeping, mood swings, and depression – which together are among the most common and distressing menopause symptoms observed in midlife women^11–17^ – plus total symptom count. A composite score of global cognition was calculated by averaging sample-based z-scores on tests for memory (Rey Auditory Verbal Learning Test total learning and delayed recall, Logical Memory IA and IIA) and executive function (Trail Making Test Part B, Weschler Adult Intelligence Scale–Digit Symbol Substitution Test, and Animal fluency). Plasma samples from a subset of recent WRAP visits (2017–2024) were analyzed for 122 proteins on the NULISAseq CNS kit.^18^ Given the lack of sufficient follow-up after the NULISAseq blood draw, we restricted analyses to cross-sectional data. Our sample included *N*=93 female participants who had cognitive data available within 30 days of NULISA blood collection. Plasma p-tau217/Aβ42 was measured on the Lumipulse G System (Fujirebio) at the Michael T. Zuendel Biomarker Laboratory at Banner Sun Health Research Institute. To characterize the sample, amyloid positivity was determined using a threshold of >0.008.^19^

***UK Biobank (UKB; aging cohort 4):*** To assess whether menopause-related proteomic shifts predict risk for dementia, we used data from *N*=11,059 women ≥50 years old in the UKB with non-missing values for all 13 menopause proteins measured on Olink at study baseline. Participants were free from known dementia at baseline and followed for a mean (*SD*) of 15.7 (2.84) years. Consistent with prior work, incident dementia cases were algorithmically defined using a combination of information in international classification of diseases (ICD) diagnosis codes in linked primary care, hospital inpatient, and death registry data, for all-cause dementia (codes A81, F00 to F03, F05, F10, G30, G31 and I67), alongside Alzheimer’s disease (F00 and G30), vascular dementia (F01 and I67) and frontotemporal dementia (F02 and G31).^5^ Participants were censored at first dementia diagnosis, death, or September 2025, representing the most recent update of algorithmically defined dementia outcomes in the UKB.

**Analyses**

**PC1 in aging cohorts:**

To accommodate varying protein availability between aging cohorts (Table S17), we recomputed several versions of PC1 in the spontaneous menopause cohort after (1) removing SNAP25 for use in ADNI; (2) removing BDNF for use in WRAP; and (3) removing BACE1, IL12p70, and p-tau231 for use in UKB. Loadings for the remaining shared proteins among the versions were very similar (Figure S9).

**Additional analytic details:**

Analyses were conducted in R (v4.5.0). Descriptive statistics and ANOVA, t-tests, and χ^2^ tests summarized demographic and clinical characteristics in each cohort. Gene ontology (GO) and pathway enrichment analyses using the publicly-available GOparallel code (<https://www.github.com/edammer/GOparallel>).^20^

**Organ aging and cell aging models:**

To assess the links between menopause timing with biological aging, in the UKB, we calculated proteomic aging clocks for 13 organ systems and 38 cell types using previously established Olink models.^5,21^ Organ and cell clock models were developed using proteins that are labeled as enriched for the corresponding tissue or cellular sources in the Human Protein Atlas (HPA),^22^ such that proteins that load onto each organ- or cell-specific model are assumed to originate at least in part from those organs/cells.^5^ Because the organ and cell clock estimations required complete data, these analyses were performed on a subset of *N*=2,340 age-matched pre/peri- and postmenopausal women following K nearest-neighbours imputation of the Olink data (below).^5^ Chronological age-adjusted linear models tested associations of age at menopause with organ and cell age “gaps” (i.e., z-scored residuals of proteomic predicted age linearly regressed against actual age, whereby larger gaps reflect more advanced biological aging).

**UK Biobank Olink imputation for organ age models:**

We imputed Olink proteomics data using the entire Olink baseline sample (*N*=52,995 participants; *N*=2,923 proteins) following a previously published method.^5^ For quality control, we first excluded *N*=8,181 samples with more than 1,000 proteins missing, seven proteins with missing values in more than 10% of samples, and 338 samples with discordant self-reported sex and genetic sex, resulting in a final dataset of *N*=44,476 participants with 2,916 protein measurements. We performed missing value imputation of the Olink data using *k*-nearest neighbors (KNN) imputation. We split the data into train and test, where each split comprised 11 randomly selected UKB assessment centers (train centers: Newcastle, Nottingham, Bristol, Leeds, Sheffield, Glasgow, Wrexham, Croydon, Hounslow, Edinburgh; test centers: Liverpool, Manchester, Middlesborough, Oxford, Stockport, Stoke, Swansea). Protein levels were z-transformed using the means and standard deviations of the train split. Using scikit-learn’s KNNImputer function (scikit-learn.org), we trained a KNN imputed where the number of neighbours (*k*) was set to the square root of the sample size in the train split (*k* = 166). We assessed the imputer’s performance on *N*=5,590 samples (3,425 train/2,165 test) with no original missing protein values. To do so, we randomly input missing values into this “ground truth” subsample at 3% (i.e., equivalent to the missing value rate in the entire post quality control dataset). We then used the imputer on this subsample to calculate the error between imputed values and ground truth values. This resulted in a total mean error rate (MAE) of 0.57 in both the train and test samples, consistent with previous research^5^ and supporting robust imputation of the Olink data (Fig.S10).

**Extended results**

**Sensitivity analyses:**

To further address the possibility that menopause proteomic differences are driven by chronological (vs. endocrine) aging, we performed sensitivity analyses repeating models in a subsample of participants with a tightly restricted age range (i.e., 47–53; *N*=54 total; *N*=17 pre-, *N*=22 peri-, *N*=15 post-menopausal). These analyses yielded very similar findings to the main results (Figure S6, Table S14), suggesting the observed proteomic differences between menopause groups were not substantially confounded by chronological age. In the UKB age-matched validation cohort, we re-ran differential abundance analyses additionally adjusting for factors that systematically differed between pre- and postmenopausal women (i.e., years of education, history of hormone therapy use, current hormonal contraceptive use, ethnicity, smoking status, LDL cholesterol, and hbA1C), which yielded highly consistent findings to the main analyses (Figure S11). In all cohorts, we also re-estimated models after adjusting for cardiometabolic comorbidities and common health-related risk factors that may influence the proteome, per available data in each cohort. These covariates included: UCSB HAS: body mass index (BMI), smoking history, and self-reported histories of high blood pressure, high cholesterol, and diabetes; ADNI: BMI, smoking history, systolic blood pressure, and diastolic blood pressure; BrANCH: BMI, smoking history, systolic blood pressure, diastolic blood pressure, hbA1c, and LDL cholesterol; WRAP: BMI, smoking history, systolic blood pressure, diastolic blood pressure, fasting glucose, and LDL cholesterol; UKB: BMI, smoking history, systolic blood pressure, diastolic blood pressure, hbA1c, and LDL cholesterol. All analyses yielded similar results to main models (Tables S15-16).

In aging cohorts, we performed additional sensitivity analyses re-estimating models after excluding participants who self-reported a history of bilateral oophorectomy (not available in ADNI) and stratified by history of hormone therapy (ever used vs. never used; not available in ADNI), which also yielded similar findings (Tables S10, S16). Additionally, given the known associations of plasma p-tau with cognitive decline and AD risk, we computed an alternate version of the menopause signature excluding p-tau231 (Figure S9). Analyses yielded a very similar pattern of results to the main analyses excluding ptau231 (Table S16). In ADNI, BrANCH, and WRAP, which had detailed information on baseline cognitive diagnoses, we also re-estimated models excluded participants with mild cognitive impairment (MCI) or dementia at baseline, which again produced similar finding (Tables S10, S16).

In both the UCSB HAS and aging cohorts, we finally performed exploratory analyses testing effect modification by *APOE4* genotype (i.e., carrier vs. non-carrier). *APOE4* carriage was inferred using NULISAseq serum levels of the APOE4 protein in the UCSB HAS cohort (Fig.S8) and was determined using genetic data for the single-nucleotide polymorphisms rs429358 and rs7412 in the aging cohorts (i.e., ADNI, BrANCH, WRAP, UKB). These showed largely null effects, with the exception that *APOE4* carriage significantly exacerbated associations of menopause proteomic scores with global cognitive decline in ADNI (Tables S15-16; Fig.S7).

**Figure S1. Variance plot for principal components analysis.**

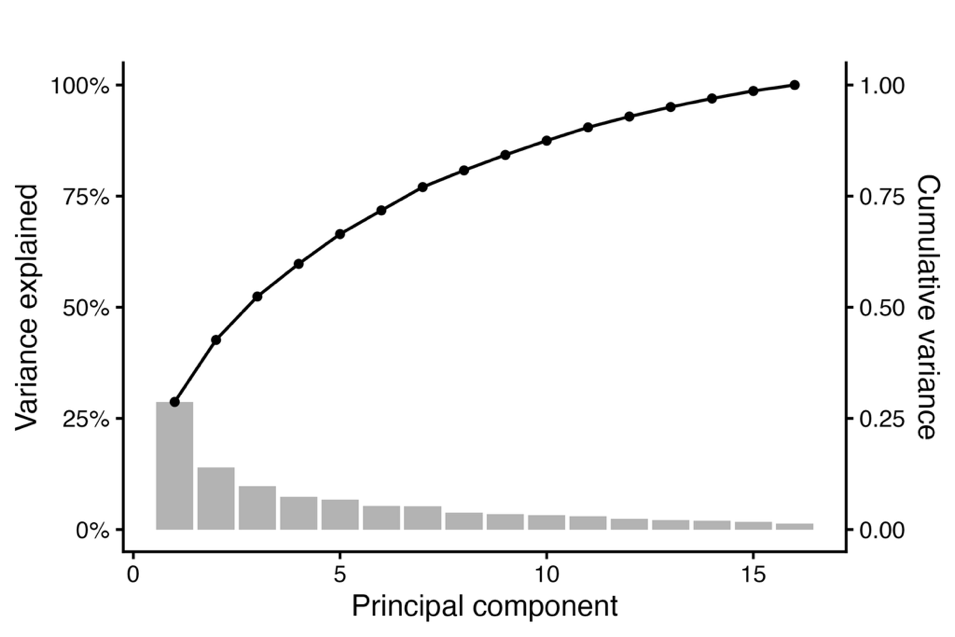

**Figure S2. Means and standard deviations (SD) of PC1 loadings across 1,000 bootstrapped resampling iterations.**

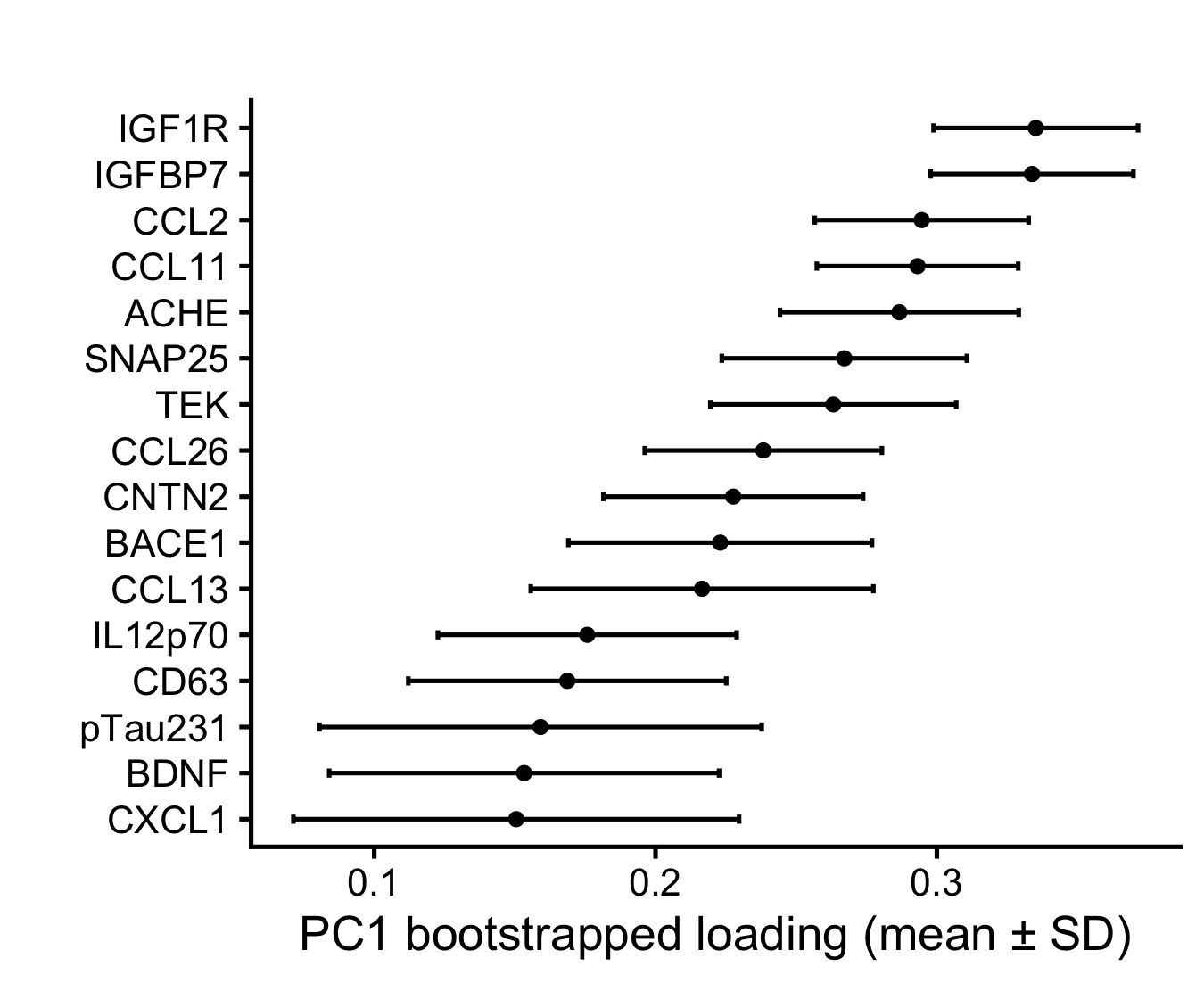

**Figure S3. Distributions of plasma biomarkers of neurodegeneration by menopause status.**

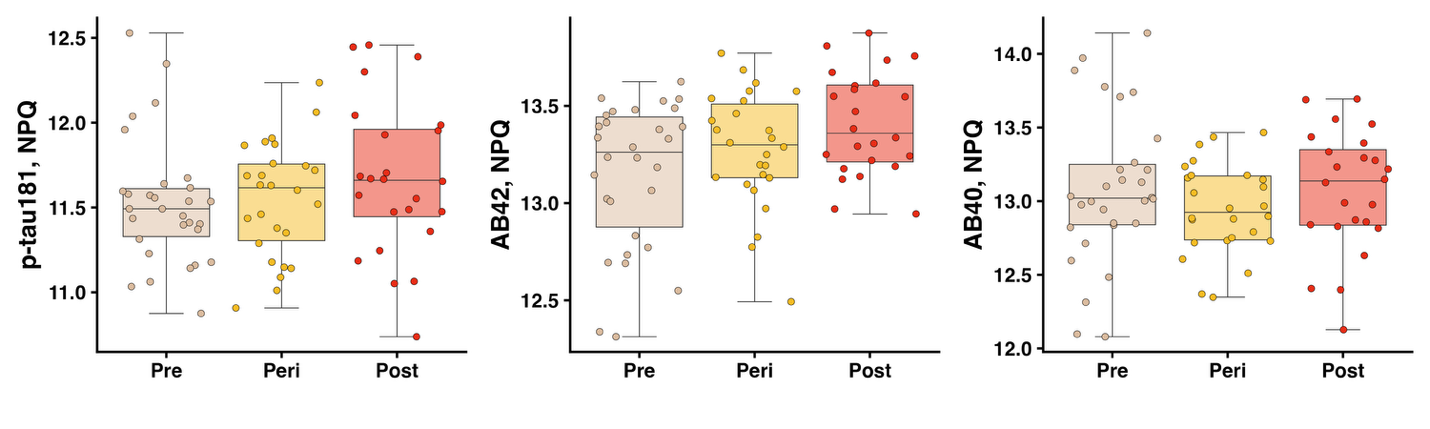

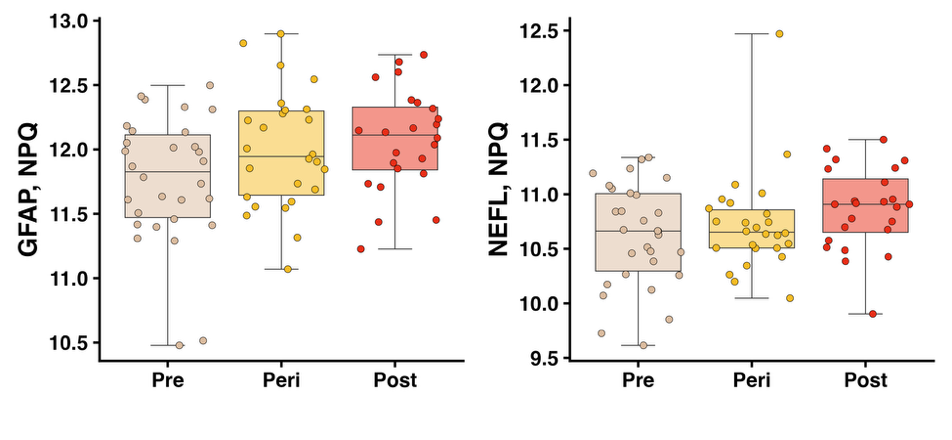

**Figure S4. Volcano plot showing organ age gaps (z-scored) by menopause stage.**

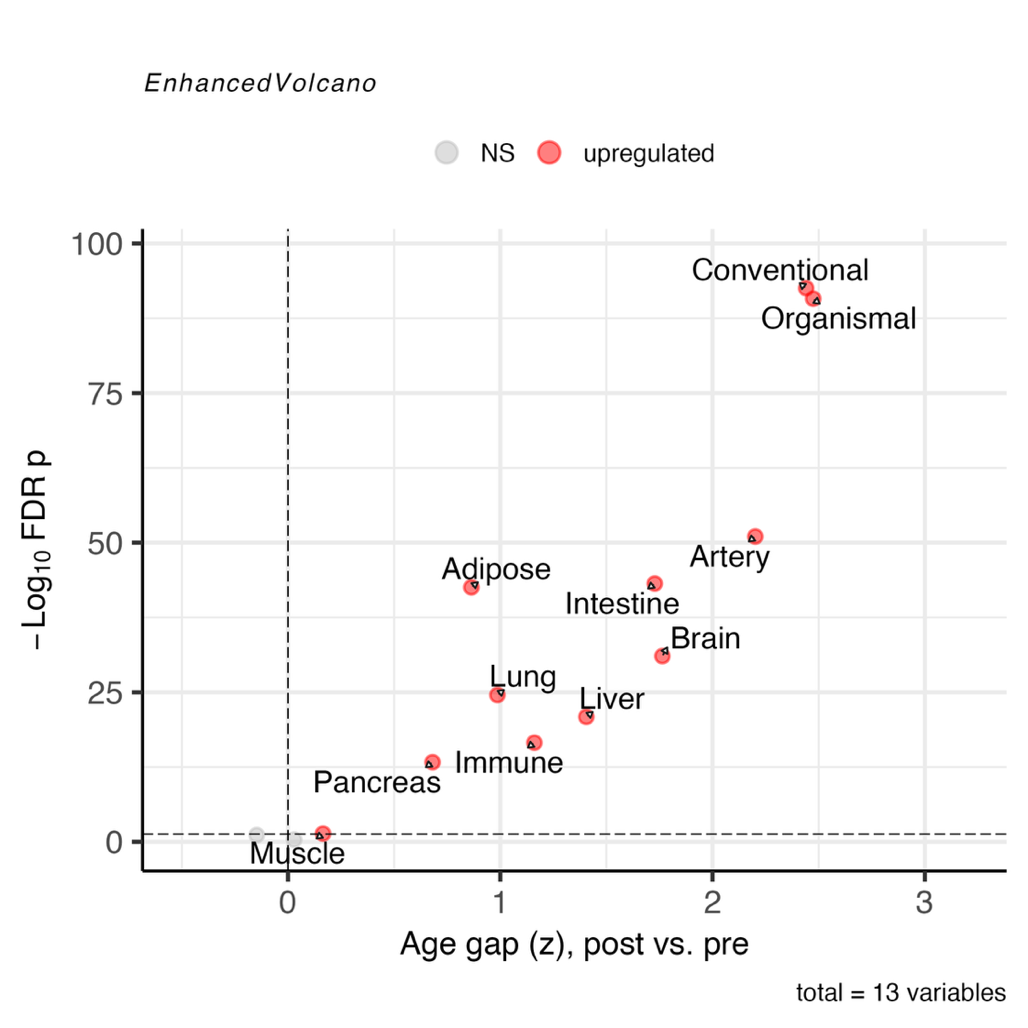

**Figure S5. Volcano plot showing cell age gaps (z-scored) by menopause stage.**

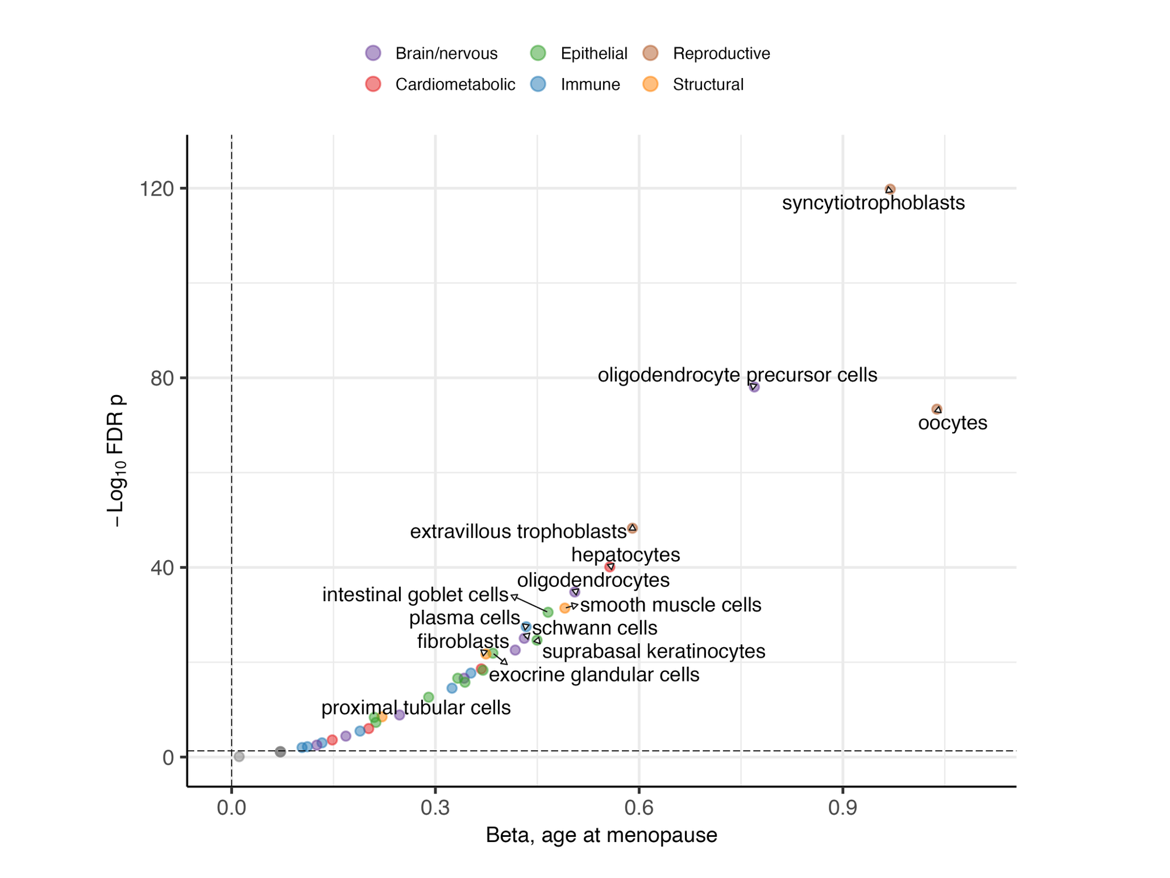

**Figure S6. Comparison of log2fold change in protein levels in postmenopausal vs. premenopausal women, in the whole UCSB analytic sample (x-axis) and an age-restricted subset (age 47-53, y-axis).**

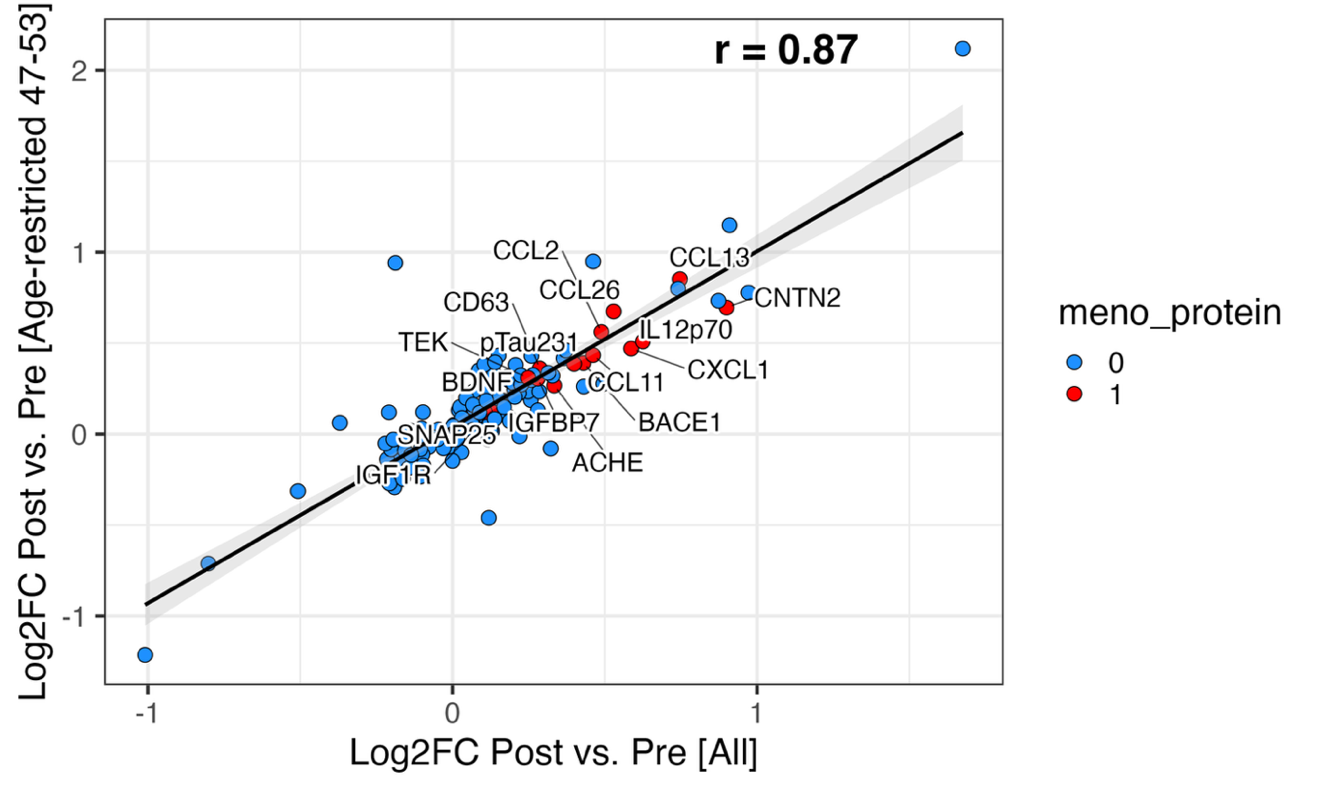

**Figure S7. Interactive associations of *APOE4* carriage and menopause proteomic scores (i.e., PC1) on global cognitive decline in ADNI.**

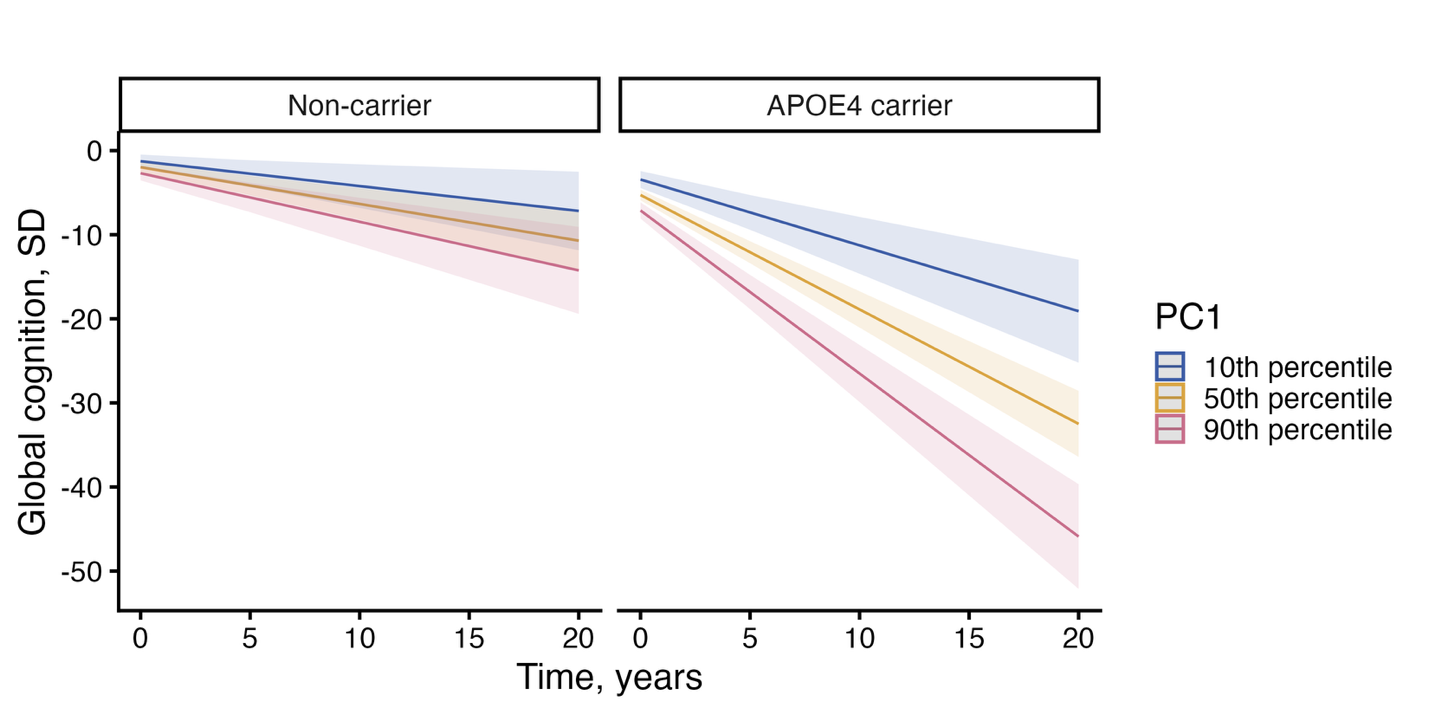

**Figure S8. Bimodal distribution of serum NULISAseq APOE4 levels used to infer *APOE4* allele carriage in UCSB HAS menopause cohort.**

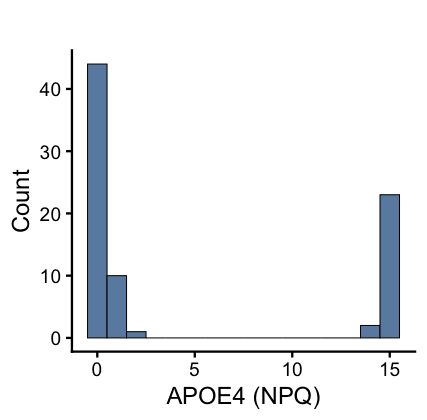

**Figure S9. Comparison of PC1 loadings with and without inclusion of BDNF, SNAP25 and p-tau231.**

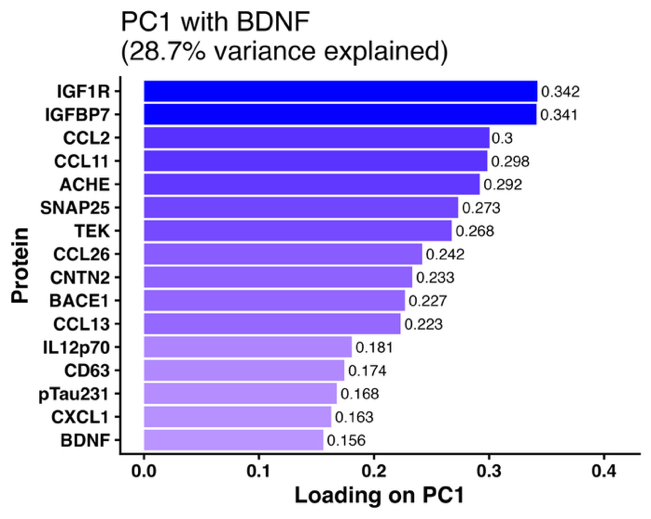

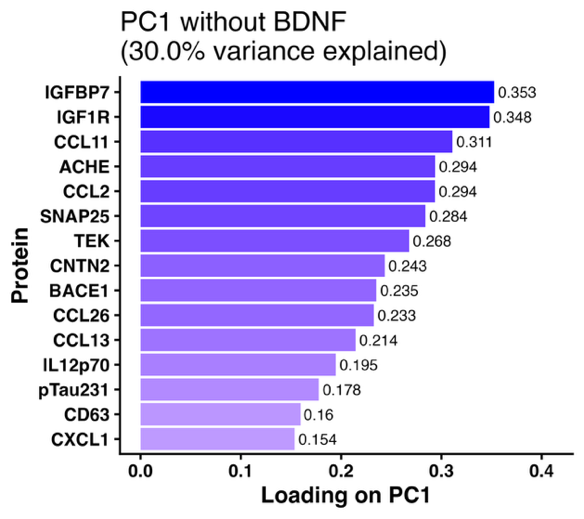

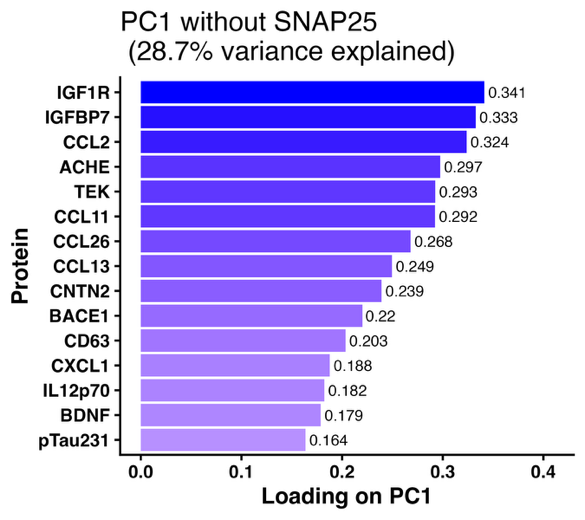

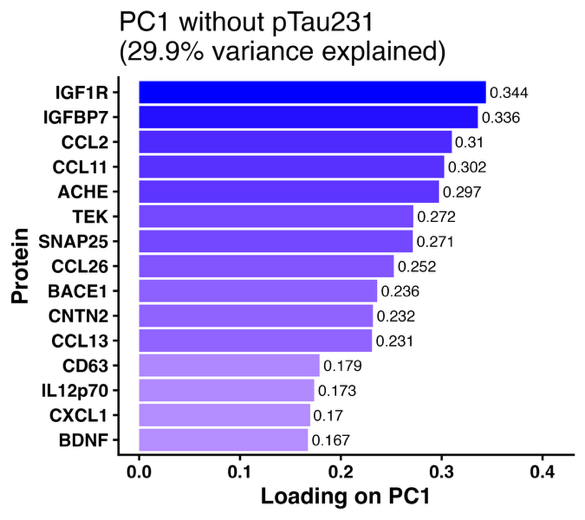

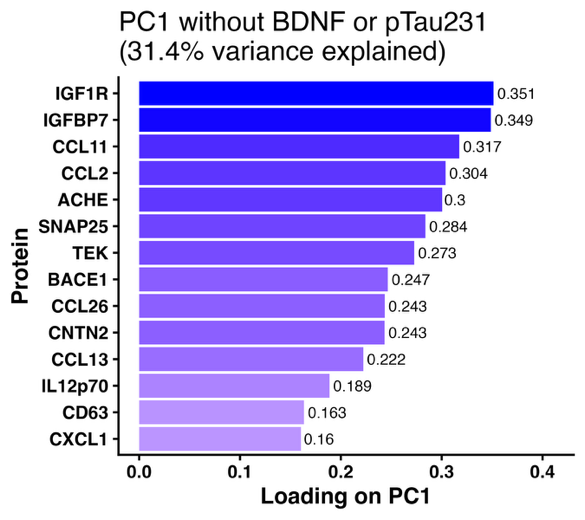

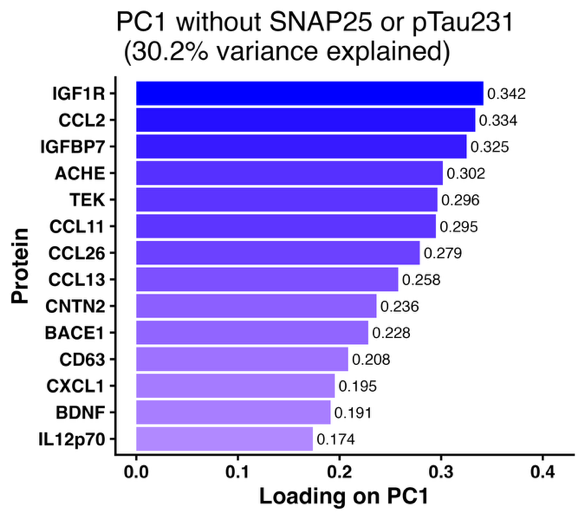

**Figure S10. Imputation evaluation metrics including distribution of ground truth vs. imputed mean average error (MAE) per protein in train and test splits (left) and total MAE rate in both train and test splits (right).**

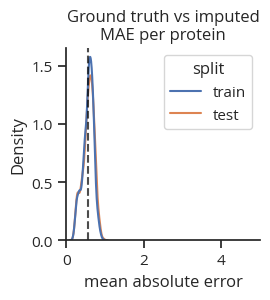

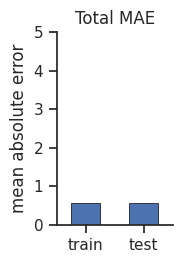

**Figure S11. Comparison of log2fold change in protein levels in postmenopausal vs. premenopausal women in the UKB, in main analyses (x-axis) and sensitivity analyses (y-axis).**

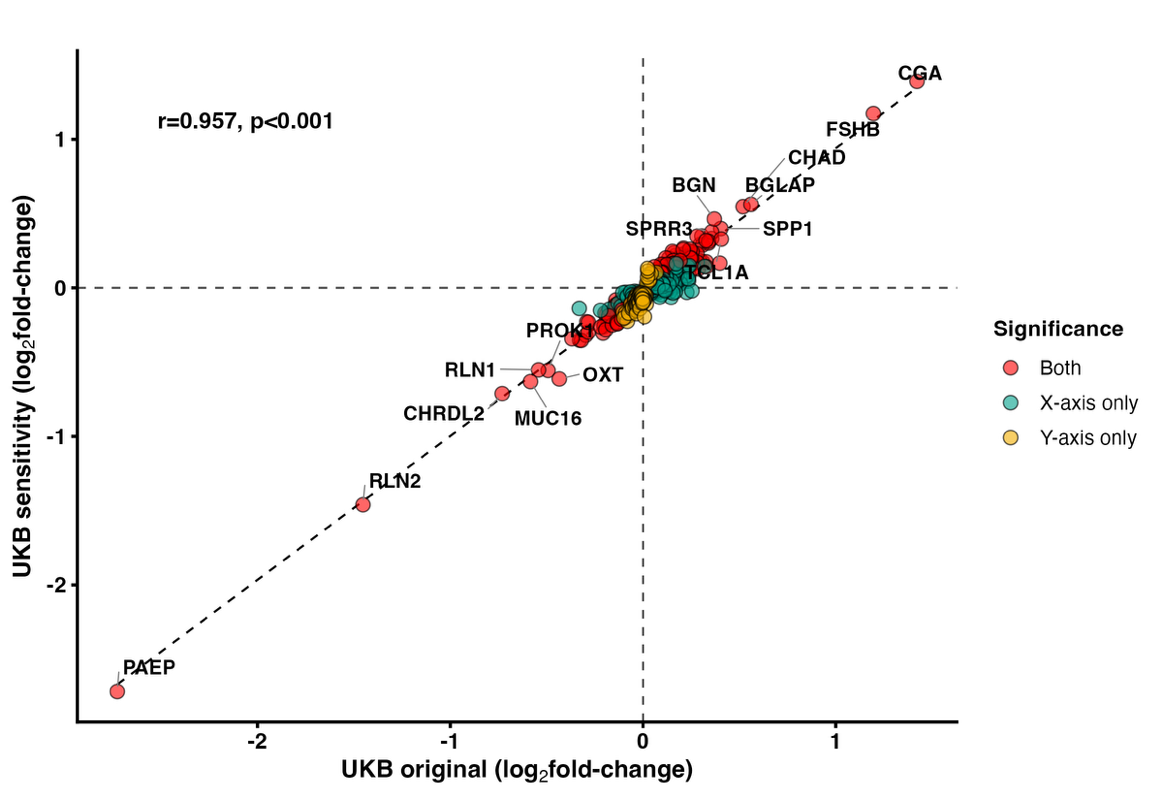

**References**

1.	Feng W, Beer JC, Hao Q, et al. NULISA: a proteomic liquid biopsy platform with attomolar sensitivity and high multiplexing. *Nat Commun*. 2023;14(1):7238. doi:10.1038/s41467-023-42834-x

2.	Zeng X, Lafferty TK, Sehrawat A, et al. Multi-analyte proteomic analysis identifies blood-based neuroinflammation, cerebrovascular and synaptic biomarkers in preclinical Alzheimer’s disease. *Mol Neurodegener*. 2024;19(1):68. doi:10.1186/s13024-024-00753-5

3.	Higginbotham L, Shantaraman A, Guo Q, et al. Multiplex Proteomics of Lewy Body Dementia Reveals Cerebrospinal Fluid Biomarkers of Disease Pathology and Progression. *BioRxiv Prepr Serv Biol*. Published online June 13, 2025:2025.06.10.658994. doi:10.1101/2025.06.10.658994

4.	Sun BB, Chiou J, Traylor M, et al. Plasma proteomic associations with genetics and health in the UK Biobank. *Nature*. 2023;622(7982):329-338. doi:10.1038/s41586-023-06592-6

5.	Oh HSH, Le Guen Y, Rappoport N, et al. Plasma proteomics links brain and immune system aging with healthspan and longevity. *Nat Med*. 2025;31(8):2703-2711. doi:10.1038/s41591-025-03798-1

6.	Petersen RC, Aisen PS, Beckett LA, et al. Alzheimer’s Disease Neuroimaging Initiative (ADNI). *Neurology*. 2010;74(3):201-209. doi:10.1212/WNL.0b013e3181cb3e25

7.	Donohue MC, Sperling RA, Petersen R, et al. Association Between Elevated Brain Amyloid and Subsequent Cognitive Decline Among Cognitively Normal Persons. *JAMA*. 2017;317(22):2305. doi:10.1001/jama.2017.6669

8.	Jagust WJ, Koeppe RA, Rabinovici GD, et al. The ADNI PET Core at 20. *Alzheimers Dement J Alzheimers Assoc*. 2024;20(10):7340-7349. doi:10.1002/alz.14165

9.	for the Alzheimer’s Disease Neuroimaging Initiative, Royse SK, Minhas DS, et al. Validation of amyloid PET positivity thresholds in centiloids: a multisite PET study approach. *Alzheimers Res Ther*. 2021;13(1):99. doi:10.1186/s13195-021-00836-1

10.	Lindbergh CA, Walker N, La Joie R, et al. Worth the Wait: Delayed Recall after 1 Week Predicts Cognitive and Medial Temporal Lobe Trajectories in Older Adults. *J Int Neuropsychol Soc*. 2021;27(4):382-388. doi:10.1017/S1355617720001009

11.	Wang Y, Islam RM, Bond M, Skiba MA, Davis SR. Sexual dysfunction in women at midlife: a cross-sectional study of data from the Australian Women’s Midlife Years study. *Lancet Obstet Gynaecol Womens Health*. 2025;1(3):e198-e208. doi:10.1016/j.lanogw.2025.100024

12.	Dennerstein L, Alexander JL, Kotz K. The menopause and sexual functioning: a review of the population-based studies. *Annu Rev Sex Res*. 2003;14:64-82.

13.	Maki PM, Panay N, Simon JA. Sleep disturbance associated with the menopause. *Menopause*. 2024;31(8):724-733. doi:10.1097/GME.0000000000002386

14.	Badawy Y, Spector A, Li Z, Desai R. The risk of depression in the menopausal stages: A systematic review and meta-analysis. *J Affect Disord*. 2024;357:126-133. doi:10.1016/j.jad.2024.04.041

15.	Maki PM, Kornstein SG, Joffe H, et al. Guidelines for the Evaluation and Treatment of Perimenopausal Depression: Summary and Recommendations. *J Womens Health*. 2019;28(2):117-134. doi:10.1089/jwh.2018.27099.mensocrec

16.	Bromberger JT, Kravitz HM. Mood and menopause: findings from the Study of Women’s Health Across the Nation (SWAN) over 10 years. *Obstet Gynecol Clin North Am*. 2011;38(3):609-625. doi:10.1016/j.ogc.2011.05.011

17.	Badawy Y, Spector A, Li Z, Desai R. The risk of depression in the menopausal stages: A systematic review and meta-analysis. *J Affect Disord*. 2024;357:126-133. doi:10.1016/j.jad.2024.04.041

18.	Rea Reyes RE, Wilson RE, Langhough RE, et al. Targeted proteomic biomarker profiling using NULISA in a cohort enriched with risk for Alzheimer’s disease and related dementias. *Alzheimers Dement J Alzheimers Assoc*. 2025;21(5):e70166. doi:10.1002/alz.70166

19.	Palmqvist S, Warmenhoven N, Anastasi F, et al. Plasma phospho-tau217 for Alzheimer’s disease diagnosis in primary and secondary care using a fully automated platform. *Nat Med*. 2025;31(6):2036-2043. doi:10.1038/s41591-025-03622-w

20.	Reimand J, Isserlin R, Voisin V, et al. Pathway enrichment analysis and visualization of omics data using g:Profiler, GSEA, Cytoscape and EnrichmentMap. *Nat Protoc*. 2019;14(2):482-517. doi:10.1038/s41596-018-0103-9

21.	Ding DY, Bot VA, Chen KL, et al. Plasma proteomic signatures of cellular aging predict human disease. *Nat Med*. Published online June 15, 2026. doi:10.1038/s41591-026-04446-y

22.	Thul PJ, Lindskog C. The human protein atlas: A spatial map of the human proteome. *Protein Sci Publ Protein Soc*. 2018;27(1):233-244. doi:10.1002/pro.3307
